# Effectiveness of a booster dose of aerosolized or intramuscular adenovirus type 5 vectored COVID-19 vaccine in adults with hybrid immunity against COVID-19: a multicenter, partially randomized, platform trial in China

**DOI:** 10.1101/2024.09.14.24313671

**Authors:** Si-Yue Jia, Yuan-Bao Liu, Qian He, Hong-Xing Pan, Zheng-Lun Liang, Juan Zhou, Ying-Zi Pan, Sheng Liu, Jing-Jing Wu, Kun Yang, Xuan-Xuan Zhang, Yang Zhao, Si-Min Li, Lei Zhang, Li Chen, Ai-Hua Yao, Meng-Yi Lu, Qun-Ying Mao, Feng-Cai Zhu, Jing-Xin Li

**Affiliations:** Jiangsu Provincial Medical Innovation Center, National Health Commission Key Laboratory of Enteric Pathogenic Microbiology, Jiangsu Provincial Center for Disease Control and Prevention (Jiangsu Provincial Academy of Preventive Medicine), Nanjing, China; Division of Hepatitis and Enterovirus Vaccines, State Key Laboratory of Drug Regulatory Science, Institute of Biological Products, National Institutes for Food and Drug Control, Beijing, China; Gaoxin (Gaogang) District Center for Disease Control and Prevention, Taizhou, China; Wujin Center for Disease Control and Prevention, Changzhou, China; Ganyu Center for Disease Control and Prevention, Lianyungang, China; Sucheng Center for Disease Control and Prevention, Suqian, China; The Affiliated Wuxi Center for Disease Control and Prevention of Nanjing Medical University, Wuxi, China; School of Public Health, National Vaccine Innovation Platform, Nanjing Medical University, Nanjing, China; School of Public Health, Southeast University, Nanjing, China

**Keywords:** COVID-19, Vaccine, Effectiveness, Adenovirus type 5, Trial

## Abstract

**Background:** The primary objective of this research was to assess if a booster dose with COVID-19 vaccines containing ancestral strain could still provide significant protection against symptomatic SARS-CoV-2 infection in a predominantly hybrid-immune population during the period of omicron variant dominance.

**Methods:** We did a multicenter, partially randomized, platform trial to evaluate the effectiveness of a booster dose of an aerosolized or intramuscular adenovirus type 5 vectored COVID-19 vaccine (Ad5-nCoV) in adults, after the national-wide omicron circulating at the end of year 2022 in China. Participants who were willing to receive a COVID-19 booster dose were randomly assigned to receive one of the booster doses. While, those participants who refused to take a booster dose but consented to participate COVID-19 surveillance were included in a control group. Both participants receiving a booster dose or not were monitored for symptomatic COVID-19 during a six-month surveillance period.

**Results:** Between May 23, 2023, and August 28, 2023, 4089 eligible participants were equally randomized to receive a booster dose of aerosolized Ad5-nCoV through oral inhalation at 0.1mL (IH Ad5-nCoV, n=2039) or intramuscular injection of Ad5-nCoV at 0.5 mL (IM Ad5-nCoV, n=2050). While, 2008 participants were enrolled in the blank-control group. A total of 79 COVID-19 cases were confirmed, with 22 (0.006%) in the IH Ad5-nCoV group, 23 (0.007%) in the IM Ad5-nCoV group, and 34 (0.01%) in the control group. Adjusted effectiveness of IH Ad5-nCoV and IM Ad5-nCoV from 14 days after the vaccination were 51.6% (95% CI 9.0 to 74.3) and 38.1% (95% CI - 9.6 to 65.1), respectively.

**Interpretation:** Significant protection against symptomatic COVID-19 caused by the Omicron variant, during the ongoing pandemic of evolving COVID-19 variants, was found to be provided by boosting with the ancestral strain-containing vaccine IH Ad5-nCoV, but not by boosting with IM Ad5-nCoV.

## Introduction

The current global population presents a complex immunological landscape regarding COVID-19, encompassing individuals who have received COVID-19 vaccinations, those who have been previously infected and recovered from the SARS-CoV-2 virus, and potentially those with hybrid immunity [1]. Variations in vaccines types administered, dosing regimens, exposure to different viral variants, and the timing of these events contribute to this intricate hybrid immune status [2]. Although a monovalent XBB.1 descendent lineage was recommended to be used as the vaccine antigen by the World Health Organization (WHO) in May 2023, they also advised that vaccination programmes of the WHO emergency-use listed or prequalified COVID-19 vaccines should not be delayed in anticipation of access to vaccines with an updated composition [3]. In addition, the scope of global vaccine inequity is immense, and its repercussions will continue to be felt worldwide, especially among the world’s most vulnerable residents [4,5]. Therefore, the question of whether booster doses of COVID-19 vaccines containing ancestral strain can still provide protection to populations with hybrid immunity during the period of omicron variant dominance remains an important area warranting investigation [6].

In China, over 90% of the population has achieved primary immunization coverage with COVID-19 vaccines, predominantly with inactivated vaccines [7]. The first national-scale outbreak of the SARS-CoV-2 omicron predominance occurred at the end of year 2022 in China, following the removal of the “dynamic zero-case policy” [8]. A survey conducted across 31 provinces in China reported that 82.4% of individuals were infected with SARS-CoV-2 between December 2022 and February 2023 [9]. Consequently, majority of the Chinese population now possesses hybrid immunity against COVID-19, derived from both vaccination with COVID-19 vaccines and breakthrough infection caused by the SARS-CoV-2 omicron variants. However, despite a significant proportion of the population acquiring a certain degree of immunity against COVID-19 through vaccination, previous infection, or a combination of both, the mutation and circulation of the SARS-CoV-2 virus continue globally, leading to the emergence of new variants [10].

Estimating effectiveness of COVID-19 booster vaccinations in a predominantly hybrid-immune population is crucial for guiding future vaccination policies aimed at controlling COVID-19. In this context, we report on the effectiveness of boost with an aerosolized or intramuscular adenovirus type 5 vectored COVID-19 vaccine (Ad5-nCoV), which has obtained emergency use authorization, in a population where the majority exhibits hybrid immunity in China.

## Methods

### Study design and participants

This is a multicenter, partially randomized, platform trial aims to evaluate the effectiveness of a booster dose of aerosolized or intramuscular adenovirus type 5 vectored COVID-19 vaccine (Ad5-nCoV) through a six-month surveillance of COVID-19 in adults aged 18 years and older with hybrid immunity against COVID-19. Participants were recruited from five cities, including Taizhou, Changzhou, Lianyungang, Suqian and Wuxi, in Jiangsu province, China. The study protocol and informed consent form were reviewed and approved by the Ethics Committee of Jiangsu Provincial Center for Disease Control and Prevention. The study was registered with ClinicalTrials.gov under the identifier NCT05855408 and conducted in accordance with the Declaration of Helsinki and Good Clinical Practice.

Eligible participants were individuals aged 18 years and older, including those over 60 years of age and individuals with underlying diseases, Eligibility required an interval ≥ 4 months after previous SARS-CoV-2 infection or confirmation of never having been infected, and ≥ 6 months since the last COVID-19 vaccination. Participants willing to receive a booster dose were randomly assigned to receive one of the boost vaccines, while those opting not to receive a booster were included as a control group. Exclusion criteria for both vaccine groups included individuals displaying suspected symptoms of COVID-19 on the day of enrollment, positive results on antigen rapid tests for SARS-CoV-2, completion of a second COVID-19 booster immunization, history of severe adverse reactions or anaphylaxis related to vaccination, and pregnant or lactating women. The exclusion criteria for the control group were the same as those for the vaccine groups, except for the absence of a history of severe adverse reactions or anaphylaxis related to vaccination. A complete list of inclusion and exclusion criteria is detailed in the study protocol. All participants in both the vaccine control groups provided written informed consent prior to screening.

### Randomization and masking

Randomization was executed using an interactive web response system (IWRS), based on a blocked randomization list generated by an independent statistician utilizing SAS software (version 9.4). Eligible participants who consented to receive a COVID-19 booster dose were randomly assigned via IWRS, to receive one of the two COVID-19 vaccines, the aerosolized Ad5-nCoV or intramuscular Ad5-nCoV, both manufactured by CanSino Biologics (Convidecia Air™ and Convidecia™). Furthermore, stratified randomization was implemented based on age categories (18-59 years or 60 years and above) and participants’ history of SARS-CoV-2 infection. The design diagram of this study is illustrated in appendix 1 p 2.

### Procedures

The aerosolized Ad5-nCoV vaccine (IH Ad5-nCoV) was orally inhaled at 0.1 mL per dose, using a continuous vaporizing system containing a nebulizer (Aerogen, Galway, Ireland) integrated by Suzhou Weiqi Biological Technology (Suzhou City, China) to aerosolise the Ad5-nCoV and generate the aerosolised droplets of vaccine into a disposable suction cup. In contrast, the intramuscular Ad5-nCoV vaccine (IM Ad5-nCoV) was administered through an intramuscularly injection of 0.5 mL. Following vaccination, all participants from both vaccine groups remained under observation at the clinic for a minimum of 30 minutes to monitor for any immediate adverse reactions.

In this study, all participants were monitored for symptom-driven COVID-19 for 6 months after the booster dose (in the vaccine groups) or from the time of enrollment (in the control group). This surveillance combined active and passive monitoring strategies. We provided antigen rapid test kits for SARS-CoV-2 infection (Vazyme, China) to both participants in the vaccine groups or control group. Participants were instructed to conduct a self-test following the product manual of the antigen rapid test kits in the event that they exhibited any COVID-19 suspected symptoms during the surveillance period. COVID-19 suspected symptoms include dry throat, sore throat, cough, fever, muscle aches, decreased or loss of smell and taste, nasal congestion, runny nose, diarrhea, conjunctivitis, fatigue, malaise, headache, dyspnea, and nausea. Participants were instructed to perform an antigen rapid test within 24 hours after the appearance of the suspected first symptom. If the initial result was negative, they were required to conduct additional tests at intervals of at least 24 hours until achieving three consecutive negative results. In cases where a positive antigen test result occurred, participants were mandated to promptly inform the investigators. Subsequently, investigators took a throat swab for the positive cases within 48 hours after receiving the positive rapid test report for nucleic acid test. Investigators followed up on each positive case through weekly telephone consultations until the resolution of symptoms or recovery. Each episode of COVID-19 was classified as mild, moderate, severe, or critical according to the grading standard issued by the National Health Commission of China [11]. Moreover, investigators conducted weekly telephone follow-ups over a 6-month period post-vaccination to monitor for serious adverse events and to remind participants about self-testing if COVID-19 suspected symptoms arose.

The immunogenicity subgroup consisted of the first 60 participants from each vaccine group. Blood samples were taken for serum and peripheral blood mononuclear cells (PBMCs) isolation before the booster dose, subsequently and at 14 days, 3 months and 6 months post-booster dose. ACE2 inhibition activities against wild-type SARS-CoV-2, B.1.1.529, BA.2.75, BA.2.75.2, BA.5, BA.4.6, BF.7, BQ.1, BQ.1.1 and XBB.1 were measured using the MSD V-Plex SARS-CoV-2 Panel 32 (ACE2) kits at a 1:50 dilution per the manufacturer’s instructions. Neutralizing antibody responses against wild-type SARS-CoV-2, XBB.1.16 and BA.4/5 were assessed using pseudovirus neutralization tests (a human immunodeficiency virus pseudovirus system expressing the spike glycoprotein). T cell immune responses were quantified using PBMCs with commercially available Human IFN-γ and IL-2 ELISpot assay kits (BD). PBMCs were stimulated with a pool of peptides spanning the SARS-CoV-2 spike protein for 20 hours at a density of 2×10^5^ cells per well. After stimulation, the plates were incubated with IFN-γ or IL-2-detecting antibodies. Spots representing IFN-γ or IL-2-producing cells were counted using an ImmunoSpot S6 Universal Reader (CTL). The final determinations were calculated by subtracting the negative stimulation background levels from the measured values. For endpoint cases of COVID-19 with cycle threshold values below 32, nucleic acid samples underwent sequencing via the next-generation sequencing technology for variant typing of SARS-CoV-2.

### Outcomes

The primary outcome was the incidence of COVID-19 endpoint cases from 14 days to 6 months after receiving the booster dose. COVID-19 endpoint cases were defined as participants with COVID-19 confirmed by positive antigen rapid test or nucleic acid test after receiving the booster dose. The secondary outcomes for effectiveness were the incidence of COVID-19 endpoint cases from 7 days and 28 days to 6 months after receiving the booster dose, and severity of COVID-19 endpoint cases. The secondary outcomes for immunogenicity included ACE2-RBD binding inhibition rates and pseudovirus neutralization antibody levels against wild-type SARS-CoV-2 and Omicron variants at 14 days, 3 months and 6 months after receiving the booster dose. Specific T-cell responses measured by enzyme-linked immunospot (ELISpot) assay were also the secondary outcomes. The secondary outcome for safety was the incidence of serious adverse events within 6 months after receiving the booster dose.

### Statistical analysis

We hypothesized that participants in the vaccinated group receiving a booster shot would exhibit approximately 50% protection compared to those in the control group who did not receive a booster shot. The cumulative incidence rate of COVID-19 endpoint cases in the control group over the 6-month period of was estimated to be around 5%, while it was projected to be about 2.5% in the vaccine group. Sample size calculation was conducted using group-sequential tests for two proportions via PASS software (version 16.0), applying a one-sided α value of 0.05 and aiming for a statistical power of 90%. This analysis indicated that each vaccine group and the control group would require a minimum of 1308 participants. Anticipating a 30% dropout rate, a target enrollment of 2000 participants per group was established.

Statistical analyses were carried out using SAS version 9.4, with all tests being two-sided at an α value of 0.05. Effectiveness analyses were done in full analysis population, which include all participants who underwent randomization and either received one dose of the vaccine or were enrolled as part of the control group). The incidence of COVID-19 in the control group was calculated based on all endpoint cases identified from the day following enrollment. Effectiveness estimates (1 - Hazard ratio) were derived using the Cox proportional hazards regression model. Moreover, effectiveness adjusted by age, sex, body mass index (BMI) and SARS-CoV-2 infection history was also calculated. Cumulative incidence data were presented using the Kaplan-Meier method. Immunogenicity analyses were restricted to the immunogenicity subgroup, comprising all participants who received vaccinations and provided blood or nasal mucosa samples after vaccination. Safety analyses were performed on the full analysis population. The χ² test or Fisher’s exact test was used for categorical data. Student’s t test was used for log-transformed antibody titers, and the Wilcoxon rank-sum test for data that were not normally distributed. The antibodies against SARS-CoV-2 were reported as geometric mean titers (GMT) with 95% CIs and the cellular responses were shown as the proportion of positive responders.

## Results

Between May 23, 2023, and August 28, 2023, a total of 4089 eligible participants were equally randomized to receive a booster dose of aerosolized Ad5-nCoV through oral inhalation at 0.1mL (IH Ad5-nCoV, n=2039) or intramuscular injection of Ad5-nCoV at 0.5 mL (IM Ad5-nCoV, n=2050). Additionally, 2008 participants were enrolled as the blank-control group. The final database lock was on March 05, 2024, post completion of a 6-month follow-up for all participants. A total of 6097 participants were included in the final analysis (figure 1). The median follow-up time for both vaccine groups and the control group was 168.0 days (IQR 168.0-180.0). The mean age of all participants was 53.1 years (SD 17.5; range 18-96), with 1810 (29.7%) individuals aged 18-44 years, and 4287 aged 45 years or older (table 1). Among the 6097 participants, 2821 (46.3%) were male, and 1569 (25.7%) reported coexisting conditions and 4555 (74.8%) reported previous SARS-CoV-2 infection histories at the enrollment. Baseline characteristics of the participants were largely similar between IH Ad5-nCoV and IM Ad5-nCoV groups, but those in both vaccine groups were significantly different from the control group (p<0.05), with younger mean age, higher proportion of female participants, higher BMI, lower proportion of participants with coexisting conditions and without SARS-CoV-2 infection history in the control group.

**Figure 1.**
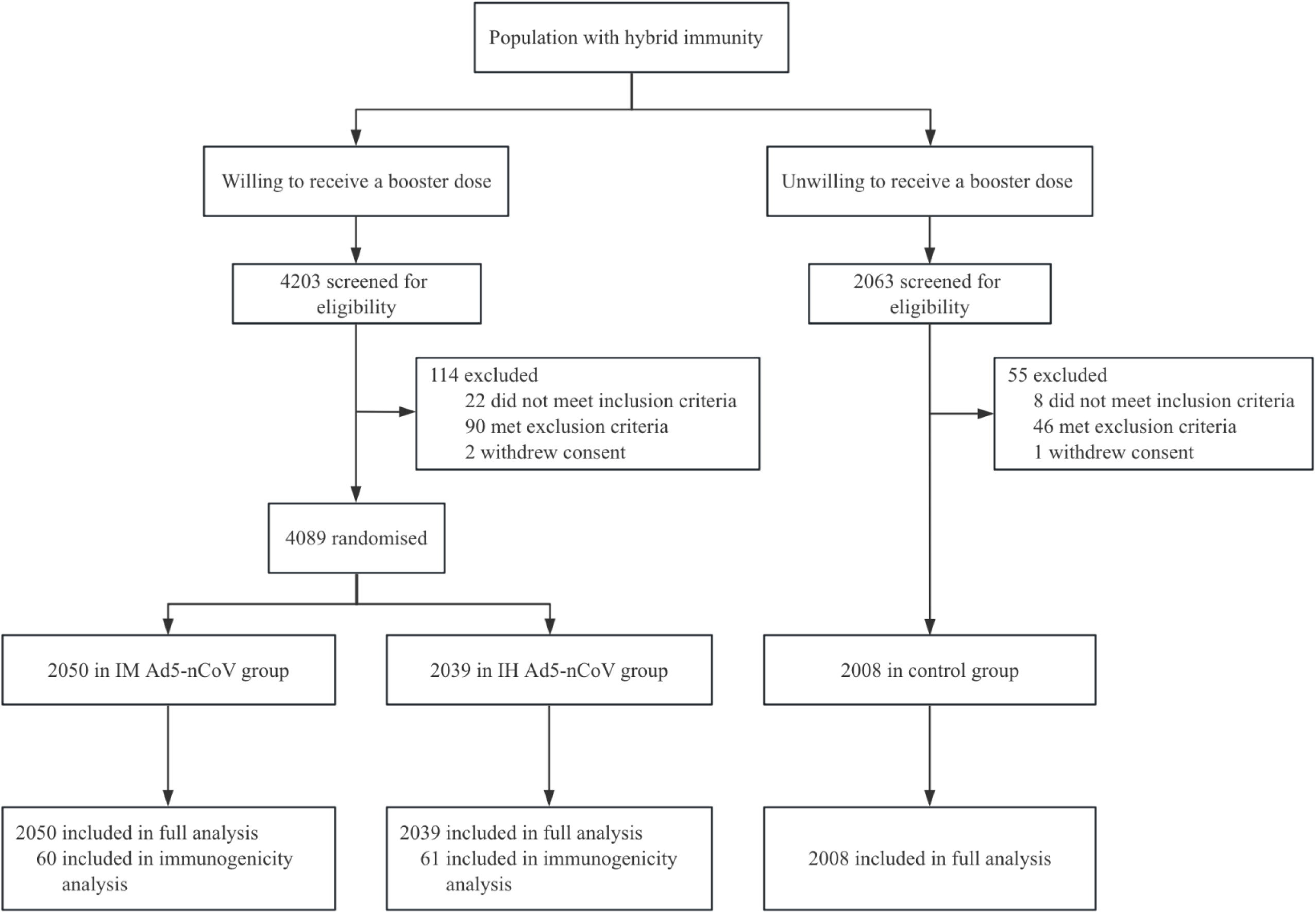
Study profile. IM Ad5-nCoV=adenovirus type 5 vectored COVID-19 vaccine through intramuscular injection. IH Ad5-nCoV= adenovirus type 5 vectored COVID-19 vaccine through oral inhalation.

**Table 1.** Demographic and baseline characteristics of the participants in full analysis population.

|  | IM Ad5-nCoV<br>(n=2050) | IH Ad5-nCoV<br>(n=2039) | Control<br>(n=2008) | IM Ad5-nCoV<br>vs. control<br>p value | IH Ad5-nCoV<br>vs. control<br>p value |
| --- | --- | --- | --- | --- | --- |
| Mean age, years | 54.5 (17.4) | 54.6 (17.3) | 50.2 (17.3) | <0.0001 | <0.0001 |
| Age, years |  |  |  |  |  |
| 18-45 | 535 (26.1) | 530 (26.0) | 745 (37.1) | <0.0001 | <0.0001 |
| ≥45 | 1515 (73.9) | 1509 (74.0) | 1263 (62.9) |  |  |
| Sex |  |  |  |  |  |
| Male | 1022 (49.9) | 998 (48.9) | 801 (39.9) | <0.0001 | <0.0001 |
| Female | 1028 (50.1) | 1041 (51.1) | 1207 (60.1) |  |  |
| BMI, kg/m² | 24.3 (3.5) | 24.3 (3.5) | 24.1 (3.6) | 0.0280 | 0.0214 |
| Coexisting conditions |  |  |  |  |  |
| Yes | 558 (27.2) | 577 (28.3) | 434 (21.6) | <0.0001 | <0.0001 |
| No | 1492 (72.8) | 1462 (71.7) | 1574 (78.4) |  |  |
| SARS-CoV-2 infection history† |  |  |  |  |  |
| Yes | 1447 (70.6) | 1451 (71.2) | 1657 (82.5) | <0.0001 | <0.0001 |
| No | 603 (29.4) | 588 (28.8) | 351 (17.5) |  |  |
| Vaccination history before booster |  |  |  |  |  |
| ICV+ICV+ICV | 1696 (82.7) | 1731 (84.9) | 1601 (79.7) | 0.0763 | 0.0003 |
| ICV+ICV | 116 (5.7) | 97 (4.8) | 143 (7.1) |  |  |
| ICV+ICV+CHO | 105 (5.1) | 98 (4.8) | 118 (5.9) |  |  |
| ICV+ICV+Ad5_IM | 59 (2.9) | 48 (2.4) | 56 (2.8) |  |  |
| CHO+CHO+CHO | 37 (1.8) | 34 (1.7) | 34 (1.7) |  |  |
| Others | 37 (1.8) | 31 (1.5) | 56 (2.8) |  |  |

Data are n (%) or mean (SD). IM Ad5-nCoV=adenovirus type 5 vectored COVID-19 vaccine through intramuscular injection. IH Ad5-nCoV= adenovirus type 5 vectored COVID-19 vaccine through oral inhalation. BMI=Body mass index.
†SARS-CoV-2 infection history was defined as having a positive antigen rapid test or nucleic acid test result or having acute onset of at least two typical symptoms or signs of COVID-19 with a history of exposure to probable or confirmed COVID-19 cases but without an etiological test result.

During the six-month surveillance, 5 confirmed COVID-19 cases in both vaccine groups (4 in the IH Ad5-nCoV group and 1 in the IM Ad5-nCoV group) were not reflected in the analysis of the primary and secondary endpoints because they occurred fewer than 7 days after the vaccination. A total of 79 COVID-19 cases were confirmed, with 22 (0.006%) in the IH Ad5-nCoV group, 23 (0.007%) in the IM Ad5-nCoV group, and 34 (0.01%) in the control group. In full analysis population, from 14 days after the vaccination, 14 COVID-19 cases were confirmed in the IH Ad5-nCoV group, 19 cases were confirmed in the IM Ad5-nCoV group, and 34 cases were confirmed in the control group, resulting in an adjusted effectiveness of IH Ad5-nCoV and IM Ad5-nCoV was 51.7% (95% CI 9.1 to 74.3) and 38.2% (95% CI -9.5 to 65.1; table 2), respectively. From 7 days after the vaccination, adjusted effectiveness of IH Ad5-nCoV and IM Ad5-nCoV was 38.8% (95% CI -9.5 to 65.7) and 29.7% (95% CI - 21.5 to 59.3). All COVID-19 cases confirmed during the surveillance period were symptomatic but mild, therefore, vaccine effectiveness against severe COVID-19 was not available. Among 29 nucleic acid samples of COVID-19 endpoint cases successfully sequenced, all were typed as Omicron variants, of which 13 (44.8%) were HK.3, 8 (27.6%) were EG.5.1.1, 3 (10.3%) were FY.3.1, 2 (10.3%) were EG.5.1, 2 (10.3%) were FY.3, and 1 (3.4%) was GF.1.

**Table 2.** Vaccine effectiveness against COVID-19 in full analysis population.

| Analysis group | Control | IM Ad5-nCoV | IH Ad5-nCoV | Unadjusted effectiveness<br>(95% CI) |  | Adjusted effectiveness*<br>(95% CI) |  |
| --- | --- | --- | --- | --- | --- | --- | --- |
|  | no. of events/follow-up in person-year (%) |  |  | IM Ad5-nCoV | IH Ad5-nCoV | IM Ad5-nCoV | IH Ad5-nCoV |
| COVID-19 occurring time |  |  |  |  |  |  |  |
| ≥ 7 days | 34/928.0 (3.7) | 22/956.9 (2.3) | 18/948.7 (1.9) | 37.0 (-7.7 to 63.1) | 47.9 (7.7 to 70.6) | 29.7 (-21.5 to 59.3) | 38.8 (-9.5 to 65.7) |
| ≥ 14 days | 34/928.0 (3.7) | 19/956.8 (2.0) | 14/948.6 (1.5) | 45.6 (4.6 to 69.0) | 59.5 (24.5 to 78.2) | 38.2 (-9.5 to 65.1) | 51.7 (9.1 to 74.3) |
| ≥ 28 days | 34/928.0 (3.7) | 14/956.5 (1.5) | 12/948.5 (1.5) | 59.9 (25.3 to 78.5) | 65.3 (32.9 to 82.0) | 53.4 (12.3 to 75.2) | 58.8 (19.6 to 78.9) |
| Subgroup analysis† |  |  |  |  |  |  |  |
| Age, years |  |  |  |  |  |  |  |
| 18-45 | 21/341.1 (6.2) | 7/248.9 (2.8) | 7/244.0 (2.9) | 53.9 (-8.4 to 80.4) | 53.2 (-10.2 to 80.1) | 58.7 (0.3 to 82.9) | 57.0 (-3.6 to 82.2) |
| ≥45 | 13/586.9 (2.2) | 12/707.9 (1.7) | 7/704.7 (1.0) | 23.5 (-67.8 to 65.1) | 55.1 (-12.6 to 82.1) | 18.4 (-79.6 to 62.9) | 51.6 (-21.5 to 80.7) |
| Coexisting conditions |  |  |  |  |  |  |  |
| Yes | 5/202.6 (2.5) | 4/259.9 (1.5) | 3/268.1 (1.1) | 37.6 (-132.3 to 83.3) | 54.8 (-89.0 to 89.2) | 37.8 (-136.6 to 83.6) | 45.6 (-134.9 to 87.4) |
| No | 29/725.4 (4.0) | 15/696.9 (2.2) | 11/680.1 (1.6) | 45.9 (-1.0 to 71.0) | 59.3 (18.4 to 79.7) | 41.9 (-9.8 to 69.3) | 54.2 (7.3 to 77.4) |
| SARS-CoV-2 infection history§ |  |  |  |  |  |  |  |
| Yes | 31/762.1 (4.1) | 15/674.9 (2.2) | 12/673.1 (1.8) | 45.1 (-1.7 to 70.4) | 55.9 (14.1 to 77.3) | 42.6 (-7.0 to 69.2) | 52.5 (7.0 to 75.8) |
| No | 3/165.9 (1.8) | 4/281.9 (1.4) | 2/275.6 (0.7) | 21.9 (-248.9 to 82.5) | 60.2 (-138.3 to 93.4) | -3.9 (-376.4 to 77.3) | 43.9 (-251.1 to 91.0) |

\*Effectiveness was adjusted by age, sex, BMI, and SARS-CoV-2 infection history.
†Subgroup analysis included COVID-19 events occurring ≥ 14 days after the vaccination.
§SARS-CoV-2 infection history was defined as having a positive antigen rapid test or nucleic acid test result or having acute onset of at least two typical symptoms or signs of COVID-19 with a history of exposure to probable or confirmed COVID-19 cases, but without an etiological test result.

Adjusted effectiveness of IH Ad5-nCoV in both age subgroups were similar (18-45 years: 57.0% [95% CI -3.6 to 82.2]; ≥45 years: 51.6% (95% CI -21.5 to 80.7) from 14 days after the vaccination. However, the protection of IM Ad5-nCoV in participants aged ≥45 years was 18.4% (95% CI -79.6 to 62.9), which was numerically much lower than the protection of 58.6% (95% CI 0.1 to 82.8) observed in those aged 18-45 years. Besides, numerically lower adjusted effectiveness of both vaccine groups was observed in participants with versus without coexisting conditions (IH Ad5-nCoV: 8.6% reduction; IM Ad5-nCoV: 4.1% reduction) and without versus with SARS-CoV-2 infection history (IH Ad5-nCoV: 8.6% reduction; IM Ad5-nCoV: 46.4% reduction; table 2, appendix 1 p 3).

As shown in figure 2, cumulative incidences of COVID-19 were similar for both vaccine groups and the control group until about 28 days after the vaccination, after which, early onset of vaccine protection led to the number of COVID-19 cases in both vaccine groups increasing much more slowly than those in the control group.

**Figure 2.**
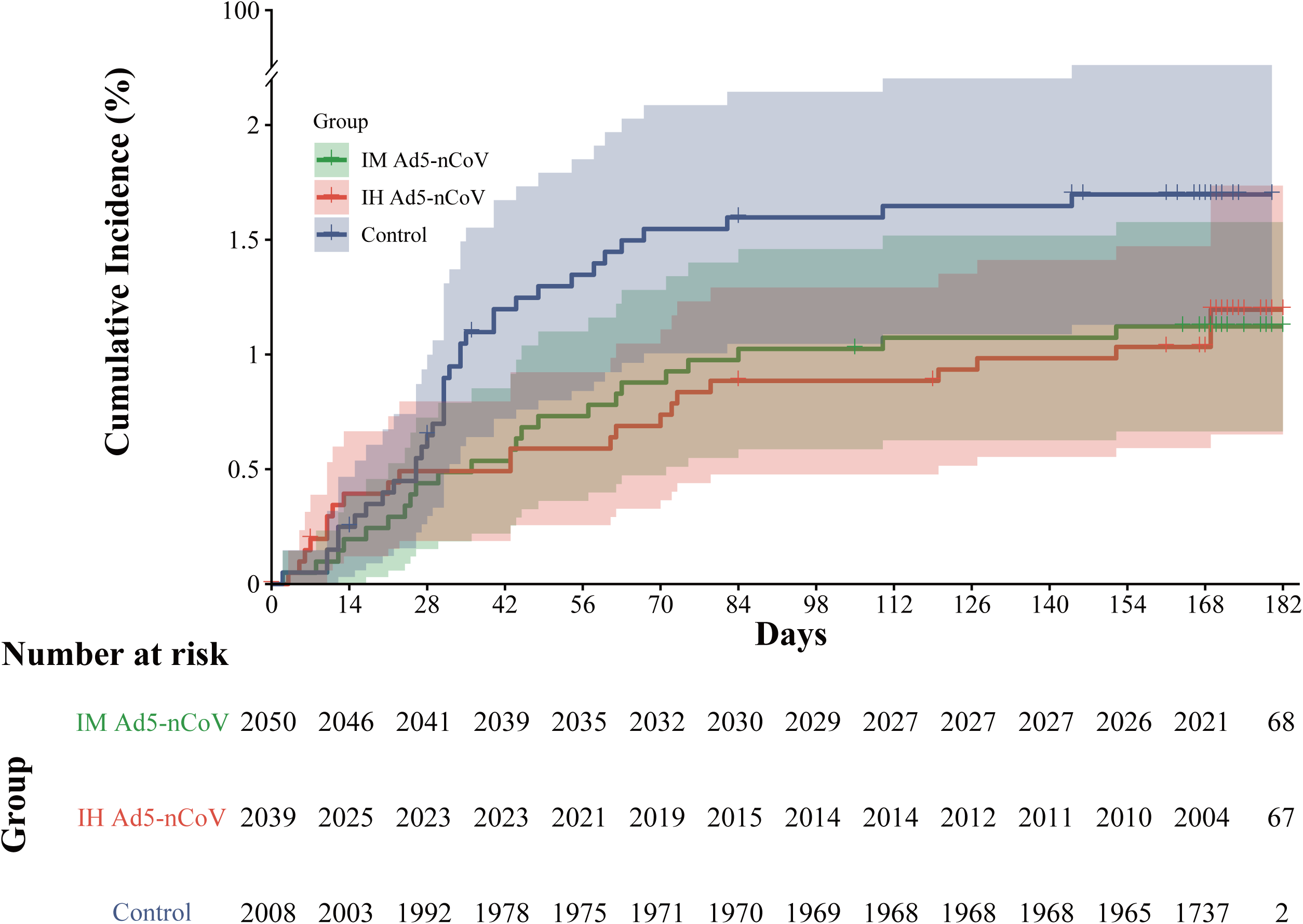
Kaplan-Meier plot of vaccine effectiveness against COVID-19 in full analysis population. IM Ad5-nCoV=adenovirus type 5 vectored COVID-19 vaccine through intramuscular injection. IH Ad5-nCoV= adenovirus type 5 vectored COVID-19 vaccine through oral inhalation. +represents participants who were censored. The shading represents the 95% confidence interval.

Immunogenicity analyses including 121 samples collected before vaccination, 14 days, 118 and 115 samples collected at months 3 and 6 post vaccination. At baseline, 65.0% of participants in IM Ad5-nCoV group and 75.4% of participants in IH Ad5-nCoV group having an inhibition of ≥50% against wild-type SARS-CoV-2; 40.0-45.0% of participants in IM Ad5-nCoV group and 50.8-59.0% of participants in IH Ad5-nCoV group having an inhibition of ≥50% against BA.4, BA.5 and BF.7 variants; but only 25.0% of participants in IM Ad5-nCoV group and 36.1% of participants in IH Ad5-nCoV group having an inhibition of ≥50% against XBB.1 variant (appendix 1 p 4). At 14 days after the booster, both vaccines elicited even higher ACE2-RBD binding inhibition against wild-type SARS-CoV-2, reaching to a proportion with an inhibition of ≥50% were 95.0% of participants in IM Ad5-nCoV group and 88.5% of participants in IH Ad5-nCoV group. At 6 months after the booster, the proportion of ACE2-RBD binding inhibition ≥50% in IM Ad5-nCoV group decreased significantly to 64.9%, but that in IH Ad5-nCoV group remained at a high level of 86.2%. Against XBB.1 variant, the proportion of ACE2-RBD binding inhibition ≥50% peaked at 14 days in IM Ad5-nCoV group, and 3 months in IH Ad5-nCoV group, which were 63.3% and 70.0%, respectively.

Pseudovirus neutralization results showed both vaccine groups have relatively high GMTs of antibody at baseline against wild-type SARS-CoV-2 (IH Ad5-nCoV: 550.4; IM Ad5-nCoV: 411.0) and BA.4/5 (IH Ad5-nCoV: 451.9; IM Ad5-nCoV: 300.9). After vaccination, pseudovirus neutralization antibody against wild-type SARS-CoV-2 in the IH Ad5-nCoV group moderately increased from 14 days and peaked at 3 months after vaccination, then slightly decreased at 6 months, with GMTs of 796.9 (95% CI 635.6 to 999.1), 1026.2 (95% CI 792.7 to 1328.6) and 880.9 (95% CI 700.9 to 1107.2), respectively (figure 3). Compared with the GMTs of pseudovirus neutralization antibody against wild-type SARS-CoV-2 in IH Ad5-nCoV group, those in IM Ad5-nCoV group was similar at 14 days, but numerically lower at 3 and 6 months, with GMTs of 796.4 (95% CI 635.3 to 998.2), 681.6 (95% CI 542.2 to 856.9) and 520.0 (95% CI 413.1 to 654.6), respectively. Compared with pseudovirus neutralization antibodies against wild-type SARS-CoV-2, those against BA.4/5 reached similar levels, with the peak GMTs at 3 months of 1061.0 (95% CI 800.1 to 1405.5) in IH Ad5-nCoV group and 883.0 (95% CI 670.1 to 1163.4) in IM Ad5-nCoV group. Although the pseudovirus neutralization antibodies against XBB.1.16 in both vaccine groups were relatively lower compared with those against wild-type and BA.4/5 SARS-CoV-2, the GMTs increased significantly after booster immunization, from 79.3 at baseline to the peak of 317.0 in IM Ad5-nCoV, and from 65.3 at baseline to the peak of 302.8, both at 3 months.

**Figure 3.**
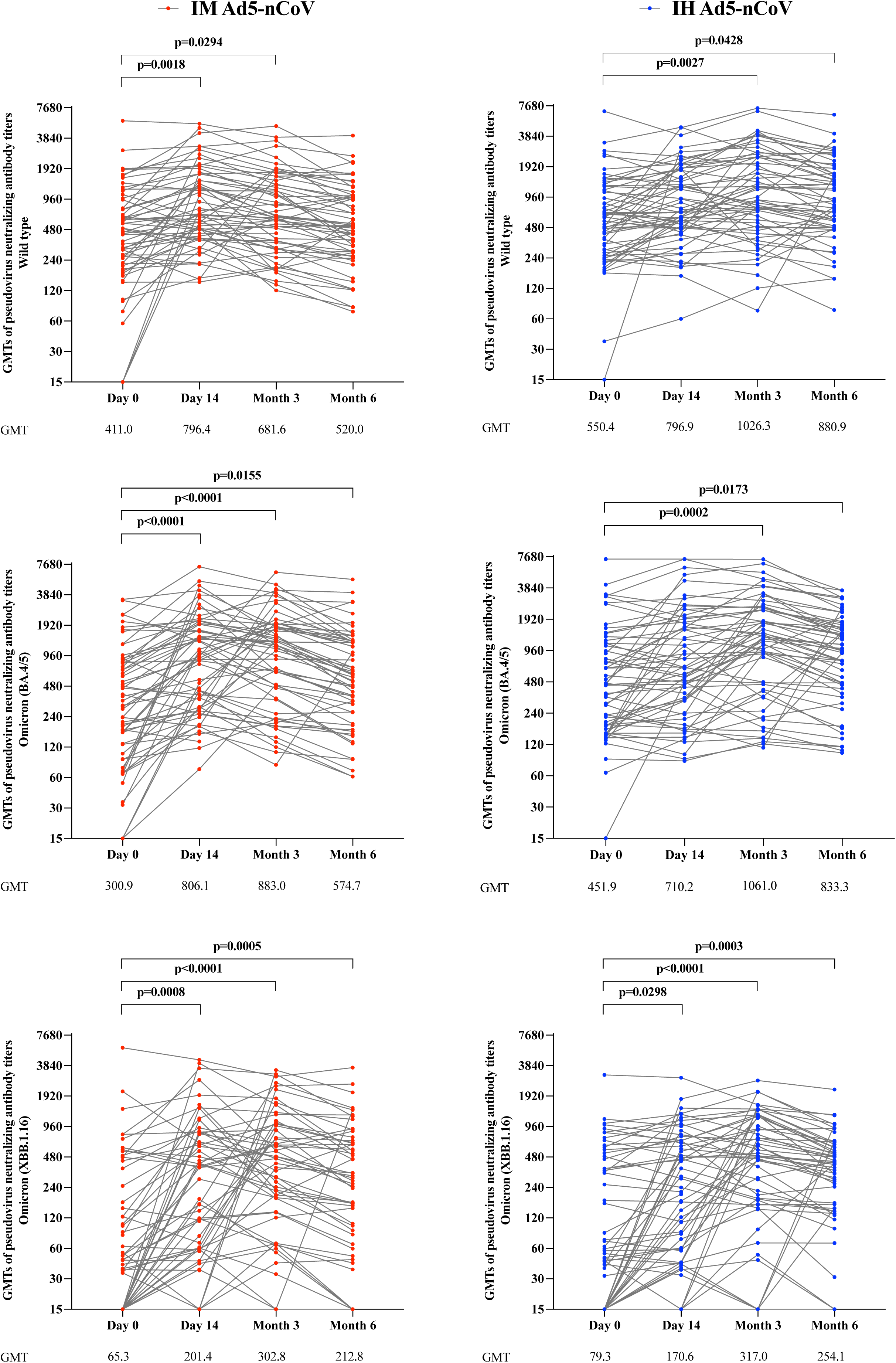
Pseudovirus neutralization antibodies against SARS-CoV-2 and variants in serum before and after a booster vaccination. IM Ad5-nCoV=adenovirus type 5 vectored COVID-19 vaccine through intramuscular injection. IH Ad5-nCoV= adenovirus type 5 vectored COVID-19 vaccine through oral inhalation. GMT=geometric mean antibody titer.

ELISpot responses were detectable before the booster dose in over 90% of participants in both vaccine groups, with a mean number of spot-forming cells per 2×10^5^ cells of 25.8 (95% CI 19.6 to 31.9) for IFN-γ and 10.2 (95% CI 7.2 to 13.1) for IL-2 in the IM Ad5-nCoV group, 27.8 (23.0-32.6) for IFN-γ and 6.7 (95% CI 5.1 to 8.6) for IL-2 in the IH Ad5-nCoV group (appendix 1 p 5). After the booster vaccination, IFN-γ and IL-2 at 14 days and 3months were similar with those at baseline, but decreased slightly at 6 months. In IH Ad5-nCoV group, the mean number of spot-forming cells per 2×10^5^ cells for IFN-γ was 22.3 (95% CI 13.5 to 31.0) at 14 days, 22.8 (95% CI 12.6 to 32.9) at 3 months, and 10.6 (95% CI 5.4 to 15.8) at 6 months, and that for IL-2 was 9.1 (95% CI 5.9 to 12.2) at 14 days, 7.9 (95% CI 5.8 to 9.9) at 3 months, and 3.7 (95% CI 0.6 to 6.6) at 6 months. In IM Ad5-nCoV group, the mean number of spot-forming cells per 2×10^5^ cells for IFN-γ was 27.3 (95% CI 15.4 to 39.1) at 14 days, 23.9 (95% CI 17.8 to 29.9) at 3 months, and 14.9 (95% CI 5.0 to 24.9) at 6 months, and that for IL-2 was 10.5 (95% CI 7.7 to 13.4 at 14 days, 8.4 (95% CI 5.9 to 10.8) at 3 months, and 4.3 (95% CI 1.9 to 6.8) at 6 months.

During a six-month follow-up period after the booster vaccination, 25 episodes of serious adverse events (SAEs) were reported in 22 participants, with in 9 (0.4%) of 2050 participants from the IM Ad5-nCoV group, 7 (0.3%) of 2039 participants from the IH Ad5-nCoV group, and 6 (0.3%) of 2008 participants from the control group (appendix 1 p 6). The incidences of SAEs between two vaccine groups and the control group were not significantly different (IM Ad5-nCoV vs. control: p=0.4618; IH Ad5-nCoV vs. control: p=0.8025). Of the 19 SAEs in both vaccine groups, the most common were respiratory thoracic and mediastinal disorders (6, 31.6%) and nervous system disorders (4, 21.1%). The one with an allergic reaction in the IH Ad5-nCoV group was considered related to the vaccination, while the others were not vaccine-related. List of all SAEs is provided in appendix 1 p 7.

## Discussion

In this trial, we found that boosting with IH Ad5-nCoV containing ancestral strain could provide significant protection against symptomatic COVID-19 associated with Omicron variants, though most of the individuals having hybrid-immune background against COVID-19 with both detectable humoral and cellular immunity at the enrollment. The adjusted effectiveness of IH Ad5-nCoV was found to be 51.7% (95% CI 9.1 to 74.3), while IM Ad5-nCoV showed lower effectiveness of 38.2% (95% CI - 9.5 to 65.1). Aligning with our findings, prior studies also reported a booster dose of mRNA COVID-19 vaccine containing ancestral strain could provide protection against the Omicron variant. A retrospective cohort study conducted in the United States revealed that a single booster dose of any mRNA COVID-19 vaccine was associated with lower risk of COVID-19 caused by the Omicron variant among individuals prime vaccinated with mRNA COVID-19 vaccines but not previously infected (hazard ratio [HR], 0.43; 95% CI, 0.41-0.46) as well as those previously infected (HR, 0.66; 95% CI, 0.58-0.76) [12]. Similarly, another prospective cohort study indicated that among previously uninfected participants who received the BNT162b2 vaccine, the adjusted vaccine effectiveness approximately 6 months post-booster was 51% (95% CI, 22 to 69) [13].

Furthermore, our findings suggest that the inhaled version of Ad5-nCoV may confer a numerically greater protection against COVID-19 compared to the intramuscular injected version of Ad5-nCoV (51.7% vs. 38.2%) but not significantly, despite similar cellular and humoral responses observed across both vaccine groups and 4/5 lower of the dosage of IH Ad5-nCoV compared to that of IM Ad5-nCoV. This is the first study to evaluate the vaccine protection offered by two different immunization routes of Ad5-nCoV on the same platform. Our results imply that individuals within the general population who possess hybrid immunity through primary vaccination and SARS-CoV-2 infection may benefit more from enhanced synergy between systemic and mucosal immunity via administration of additional mucosal vaccines. Previous studies have indicated that IH Ad5-nCoV induces more robust mucosal IgA responses compared to IM Ad5-nCoV in inactivated COVID-19 vaccines recipients [14,15], this could explain the higher effectiveness of IH Ad5-nCoV in population with established hybrid immunity against COVID-19.

Moreover, we observed a numerically decreased effectiveness of both vaccines in participants aged ≥45 years, or those with coexisting conditions. Even so, the adjusted effectiveness for IH Ad5-nCoV in those aged ≥45 years and with coexisting conditions was still 51.6% (95% CI -21.5 to 80.7) and 45.6% (95% CI -134.9 to 87.4), this finding supports the necessity for a booster dose among individuals at high risk of being infected with omicron variants, and aligns with the recommendations on COVID-19 vaccination updated by World Health Organization’s Strategic Advisory Group on Immunization [16]. Notably, numerically lower effectiveness was observed in both vaccine groups among participants without versus with SARS-CoV-2 infection history, with 8.6% and 46.4% reductions in IH Ad5-nCoV group and IM Ad5-nCoV group, respectively. This observation corresponds with literature indicating that hybrid immunity engenders stronger and broader immune responses, with high-quality memory B cells generated at 5- to 10-fold higher levels, versus infection or vaccination alone and protection against symptomatic disease lasting for 6-8 months [17,18]. However, caution must be exercised in interpreting this finding due to the relatively small proportion of participants lacking a history of SARS-CoV-2 infection. Results from ACE2-RBD binding inhibition and pseudovirus neutralization assays indicated that most participants in both vaccine groups had immune responses against wild-type SARS-CoV-2 at baseline, as well as BA.5, BF.7 and BA.4 variants. This is consistent with the background that majority of Chinese population had received COVID-19 vaccines targeting the wild-type strain, and the predominant circulation of Omicron BA.5.2 and BF.7 at the end of 2022 in China [19]. After the booster vaccination, ACE2-RBD binding inhibition and pseudovirus neutralization levels in IM Ad5-nCoV group swiftly peaked at mainly 14 days after boosting, while those in IH Ad5-nCoV group peaked at 3 months with a relative lower rising speed in our study, which is in line with that reported in a previous phase 4 trial evaluating the immunogenicity of a second booster of IH Ad5-nCoV or IM Ad5-nCoV following three doses of CoronaVac in China [15]. Although the booster vaccines used in our study were based on the ancestral strain of SARS-CoV-2, favourable cross-neutralizing against several Omicron subvariants after booster vaccination were observed, including B.1.1.529, BA.2.75, BA.2.75.2, BA.5, BA.4.6, BF.7, BQ.1, BQ.1.1 and XBB.1.

Despite the significant addition of protection and humoral responses elicited by the booster dose of IH Ad5-nCoV or IM Ad5-nCoV, no obvious T-cell responses induced after the boosting were noted in our study. One reason for this is that the relative high-level T-cell responses at baseline may hamper the further increase of cellular responses. In a previously reported study, sustained T-cell immunity was also found in COVID-19 patients at 7 months post-infection [20].

In May 2023, the WHO Technical Advisory Grouzp on COVID-19 Vaccine Composition (TAG-CO-VAC) recommended the use of a monovalent XBB.1 descendent lineage, such as XBB.1.5, as the vaccine antigen [3]. Since then, several COVID-19 vaccines with a monovalent XBB.1.5 formulation have been approved for use by regulatory authorities and introduced into COVID-19 vaccination programmes in some countries, including mRNA, protein-based and viral vector vaccines [21,22]. As of April 2024, the TAG-CO-VAC advised the use of a monovalent JN.1 lineage as the antigen in future formulations of COVID-19 vaccines [23]. However, in accordance with WHO SAGE policy, vaccination programmes should continue to use any of the WHO emergency-use listed or prequalified COVID-19 vaccines and vaccination should not be delayed in anticipation of access to vaccines with an updated composition. Furthermore, vaccine prices and cold chain systems have reduced the accessibility of COVID-19 vaccines in low/middle-income countries, let alone the recently updated monovalent COVID-19 vaccines containing XBB.1 variant [24]. Our study provide evidence for the IH Ad5-nCoV containing ancestral strain as an alternative option when the updated COVID-19 vaccine containing a new variant is not available.

However, there are some limitations of our study. First, all COVID-19 endpoint cases in our study were driven by symptoms, the protection of a booster dose of IH Ad5-nCoV or IM Ad5-nCoV against asymptomatic SARS-COV-2 infection were not obtained. Second, the severity of all COVID-19 endpoint cases founded in our trial were mild, the effectiveness of both vaccines against severe disease or hospital admission could not be evaluated. Third, participants of the blank control group in our trial were not randomized, which may have led to some confounding bias in baseline characteristics between the vaccine groups and the control group, thus the effectiveness adjusted by age, sex, BMI, and SARS-CoV-2 infection history was also calculated. Fourth, the absence of data on mucosal IgA responses restricts us to fully compare the immunogenicity of IH Ad5-nCoV and IM Ad5-nCoV, which deserves further exploration in future studies. Finally, both vaccines evaluated in this study were designed based on wild type SARS-CoV-2, because the COVID-19 vaccines containing XBB.1 variant were not available in China prior to the initiation of this trial, and the effectiveness of the two vaccines was only observed for 6 months, long-term protection is not yet available.

In conclusion, our study showed that significant protection against symptomatic COVID-19 caused by the Omicron variant, during the ongoing pandemic of evolving COVID-19 variants, was found to be provided by boosting with the ancestral strain-containing vaccine IH Ad5-nCoV, but not by boosting with IM Ad5-nCoV.

## Supporting information

CONSORT 2010 Checklist

## Data Availability

All data produced in the present study are available upon reasonable request to the authors

## Contributors

S-YJ, Y-BL, and QH were co-first authors. J-XL, F-CZ, and Q-YM were joint corresponding authors. J-XL is the principal investigator of this trial. J-XL, F-CZ, and Y-BL designed the trial and the study protocol. S-YJ drafted the manuscript. QH, Z-LL, X-XZ, and Q-YM were responsible for laboratory analyses. H-XP, LZ, F-CZ, and J-XL contributed to the critical review and revision of the article. H-XP and LZ contributed to the data interpretation. JZ, Y-ZP, SL, J-JW, and KY led and participated in the site work, including recruitment, follow-up, and data collection. YZ and M-YL were responsible for the statistical analysis. S-ML, LC, and A-HY contributed to the literature search.

## Data sharing

The study protocol is available in Supplementary appendix 2. Researchers who provide a scientifically sound proposal are allowed access to the de-identified individual participant data. Individual participant data can be obtained with a request to the corresponding authors.

## Acknowledgments

We thank CanSino Biologics for providing the investigational vaccines for this study. We also thank National Key Research and Development Program of China (grant number 2023YFC2307601), National Natural Science Foundation of China (grant numbers 82341031, 82173584, and 82222062), Major Research Plan of the National Natural Science Foundation of China (grant number 92269205), Jiangsu Provincial Science Fund for Distinguished Young Scholars (grant number BK20220064), Jiangsu Provincial Key Project of Science and Technology Plan (grant numbers BE2021738 and BE2023601), and Fund of State Key Laboratory of Drug Regulatory Science (grant number 2023SKLDRS0114) for their funding contributions.

