## Appendix 1 for "Effectiveness of a booster dose of aerosolized or intramuscular adenovirus type 5 vectored COVID-19 vaccine in adults with hybrid immunity against COVID-19: a multicenter, partially randomized, platform trial in China"

### SUPPLEMENTARY APPENDIX

**Figure S1: Design diagram**

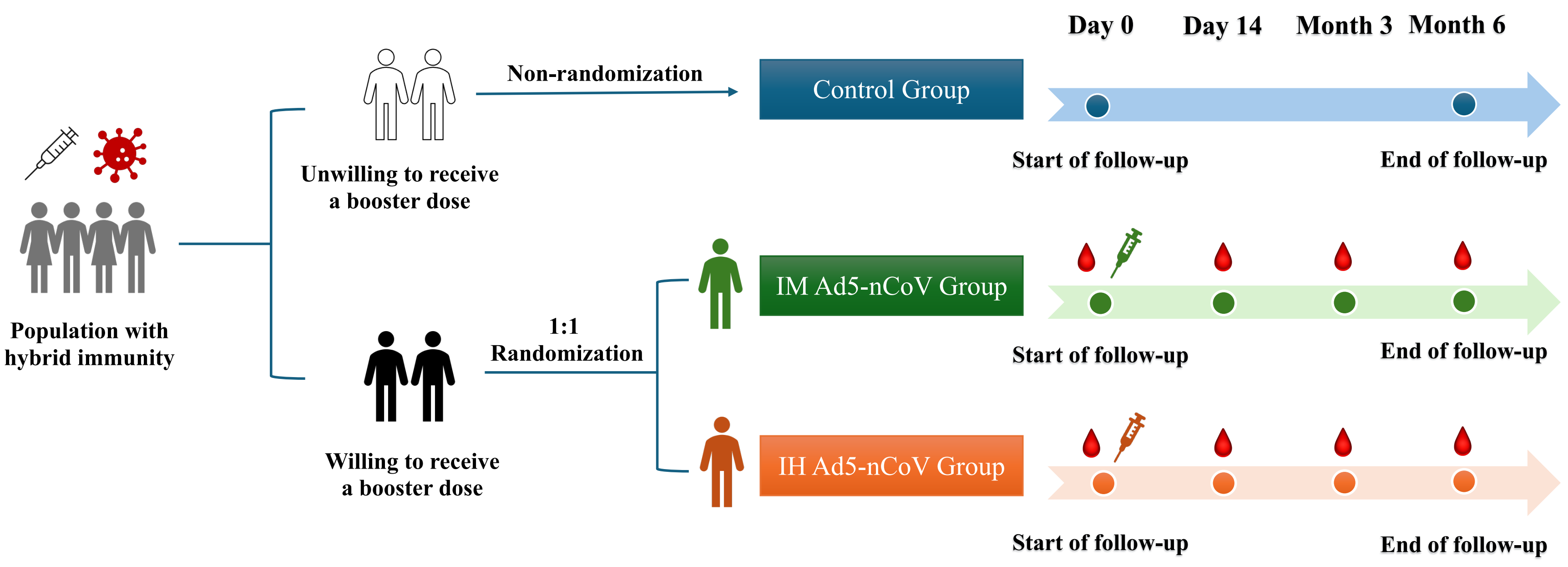

#### Figure S2: Kaplan-Meier plot of vaccine effectiveness against COVID-19 according to subgroups in full analysis population

##### A Participants aged 18−45 years

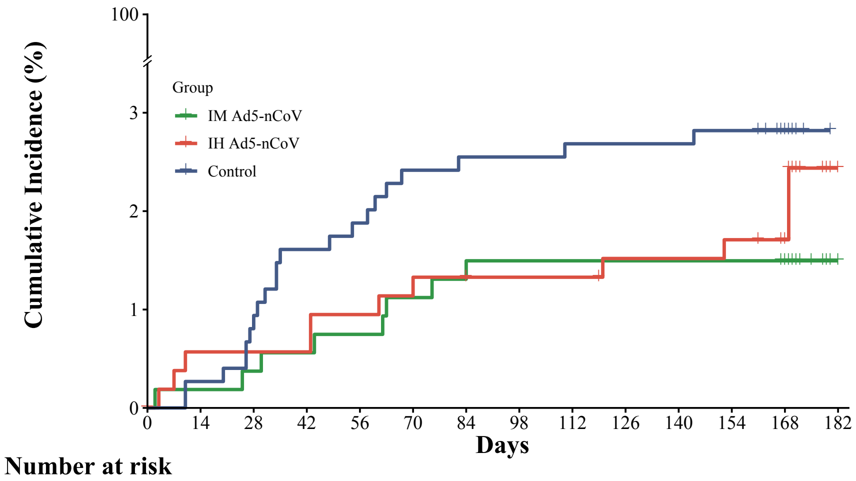

Number at risk

| Group | 0 | 14 | 28 | 42 | 56 | 70 | 84 | 98 | 112 | 126 | 140 | 154 | 168 | 182 |
|---|---|---|---|---|---|---|---|---|---|---|---|---|---|---|
| IM Ad5-nCoV | 535 | 534 | 533 | 532 | 531 | 529 | 528 | 527 | 527 | 527 | 527 | 527 | 525 | 34 |
| IH Ad5-nCoV | 530 | 524 | 524 | 524 | 522 | 521 | 520 | 519 | 519 | 517 | 517 | 516 | 511 | 24 |
| Control | 745 | 743 | 739 | 733 | 731 | 727 | 726 | 726 | 725 | 725 | 725 | 724 | 599 | 2 |

##### B Participants aged ≥ 45 years

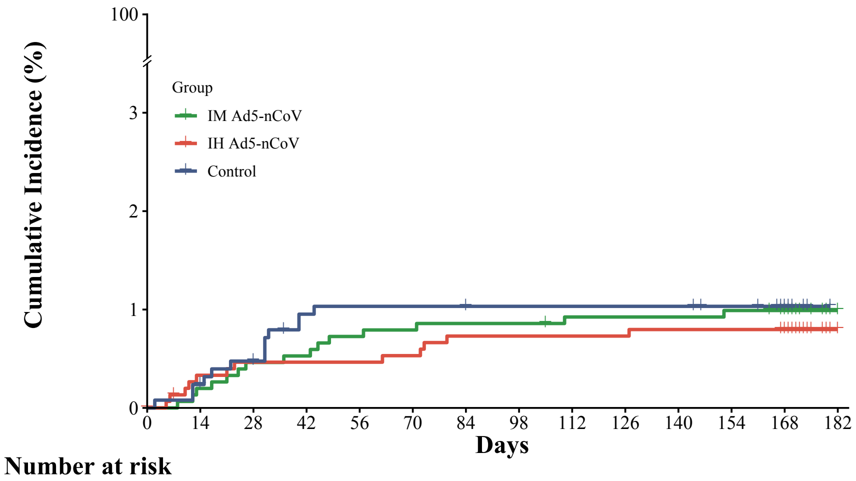

Number at risk

| Group | 0 | 14 | 28 | 42 | 56 | 70 | 84 | 98 | 112 | 126 | 140 | 154 | 168 | 182 |
|---|---|---|---|---|---|---|---|---|---|---|---|---|---|---|
| IM Ad5-nCoV | 1515 | 1512 | 1508 | 1507 | 1504 | 1503 | 1502 | 1502 | 1500 | 1500 | 1500 | 1499 | 1496 | 34 |
| IH Ad5-nCoV | 1509 | 1501 | 1499 | 1499 | 1499 | 1498 | 1495 | 1495 | 1495 | 1495 | 1494 | 1494 | 1493 | 43 |
| Control | 1263 | 1260 | 1253 | 1245 | 1244 | 1244 | 1244 | 1243 | 1243 | 1243 | 1243 | 1241 | 1138 | 0 |

##### C Participants with medical history

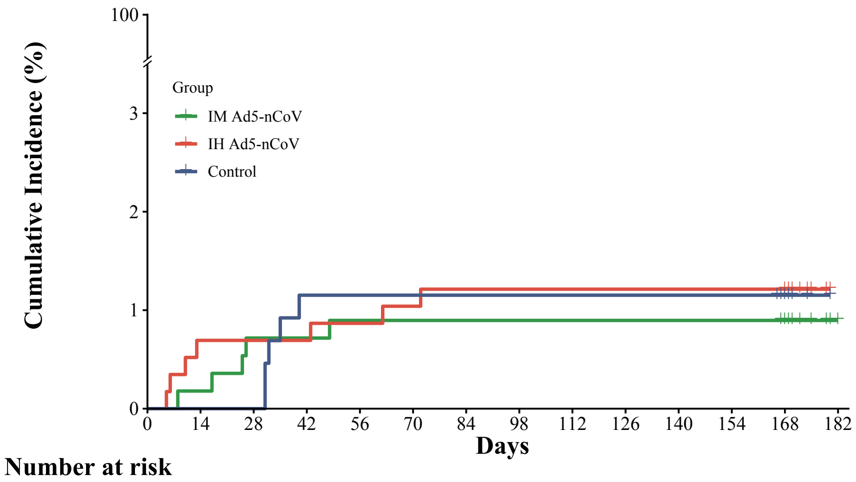

Number at risk

| Group | 0 | 14 | 28 | 42 | 56 | 70 | 84 | 98 | 112 | 126 | 140 | 154 | 168 | 182 |
|---|---|---|---|---|---|---|---|---|---|---|---|---|---|---|
| IM Ad5-nCoV | 558 | 557 | 554 | 554 | 553 | 553 | 553 | 553 | 553 | 553 | 553 | 553 | 552 | 18 |
| IH Ad5-nCoV | 577 | 573 | 573 | 573 | 572 | 571 | 570 | 570 | 570 | 570 | 570 | 570 | 570 | 15 |
| Control | 434 | 434 | 434 | 429 | 429 | 429 | 429 | 429 | 429 | 429 | 429 | 429 | 405 | 0 |

##### D Participants without medical history

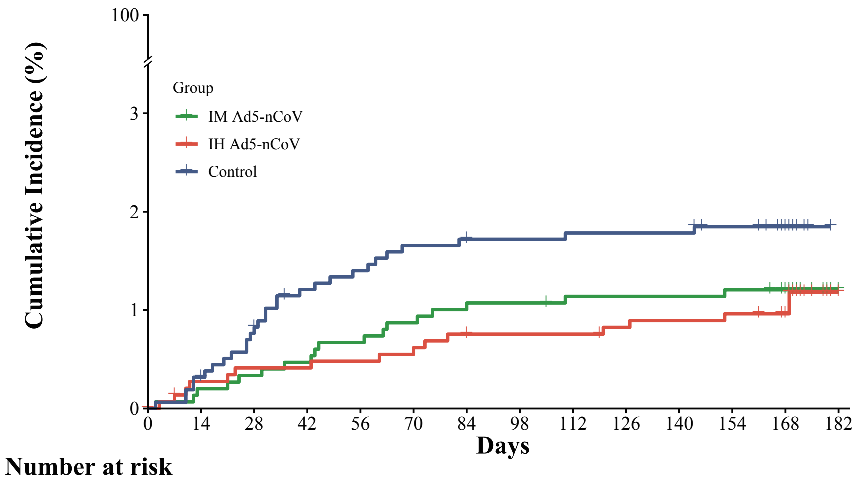

Number at risk

| Group | 0 | 14 | 28 | 42 | 56 | 70 | 84 | 98 | 112 | 126 | 140 | 154 | 168 | 182 |
|---|---|---|---|---|---|---|---|---|---|---|---|---|---|---|
| IM Ad5-nCoV | 1492 | 1489 | 1487 | 1485 | 1482 | 1479 | 1477 | 1476 | 1474 | 1474 | 1474 | 1473 | 1469 | 50 |
| IH Ad5-nCoV | 1461 | 1451 | 1449 | 1449 | 1448 | 1447 | 1444 | 1443 | 1443 | 1441 | 1440 | 1439 | 1433 | 52 |
| Control | 1574 | 1569 | 1558 | 1549 | 1546 | 1542 | 1541 | 1540 | 1539 | 1539 | 1539 | 1536 | 1332 | 2 |

##### E Participants with SARS−CoV−2 infection history

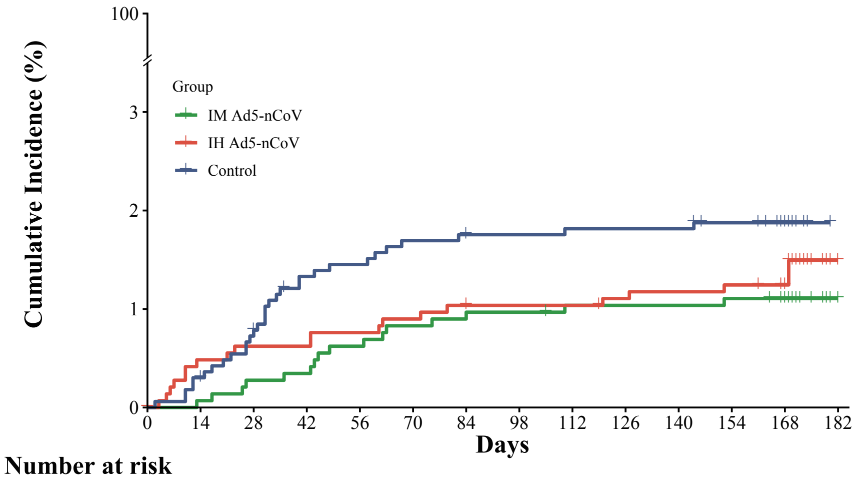

Number at risk

| Group | 0 | 14 | 28 | 42 | 56 | 70 | 84 | 98 | 112 | 126 | 140 | 154 | 168 | 182 |
|---|---|---|---|---|---|---|---|---|---|---|---|---|---|---|
| IM Ad5-nCoV | 1447 | 1446 | 1443 | 1442 | 1438 | 1435 | 1434 | 1433 | 1431 | 1431 | 1431 | 1430 | 1426 | 61 |
| IH Ad5-nCoV | 1451 | 1440 | 1438 | 1438 | 1436 | 1434 | 1432 | 1431 | 1431 | 1429 | 1428 | 1427 | 1422 | 59 |
| Control | 1657 | 1652 | 1641 | 1629 | 1627 | 1623 | 1622 | 1621 | 1620 | 1620 | 1620 | 1617 | 1431 | 2 |

##### F Participants without SARS−CoV−2 infection history

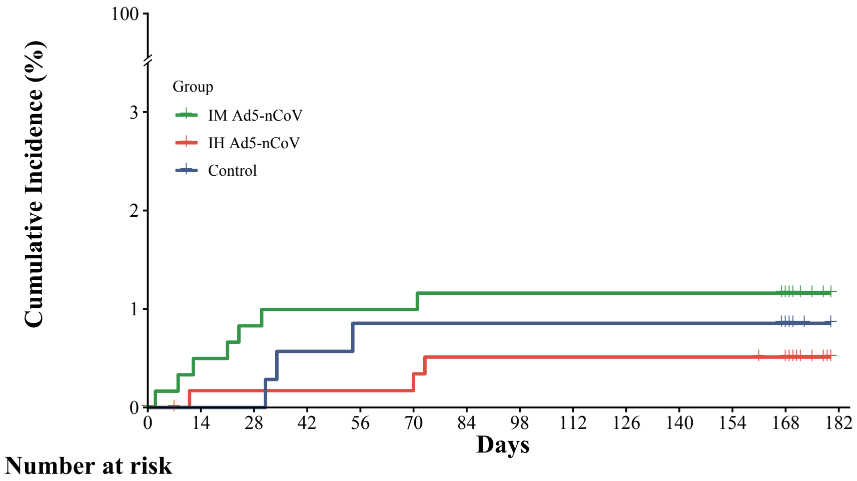

Number at risk

| Group | 0 | 14 | 28 | 42 | 56 | 70 | 84 | 98 | 112 | 126 | 140 | 154 | 168 | 182 |
|---|---|---|---|---|---|---|---|---|---|---|---|---|---|---|
| IM Ad5-nCoV | 603 | 600 | 598 | 597 | 597 | 597 | 596 | 596 | 596 | 596 | 596 | 596 | 595 | 7 |
| IH Ad5-nCoV | 588 | 585 | 585 | 585 | 585 | 585 | 583 | 583 | 583 | 583 | 583 | 583 | 582 | 8 |
| Control | 351 | 351 | 351 | 349 | 348 | 348 | 348 | 348 | 348 | 348 | 348 | 348 | 306 | 0 |

+ represents participants who were censored

**Figure S3: ACE2-RBD binding inhibition (%) against spikes of SARS-CoV-2 and variants in serum before and after a booster vaccination**

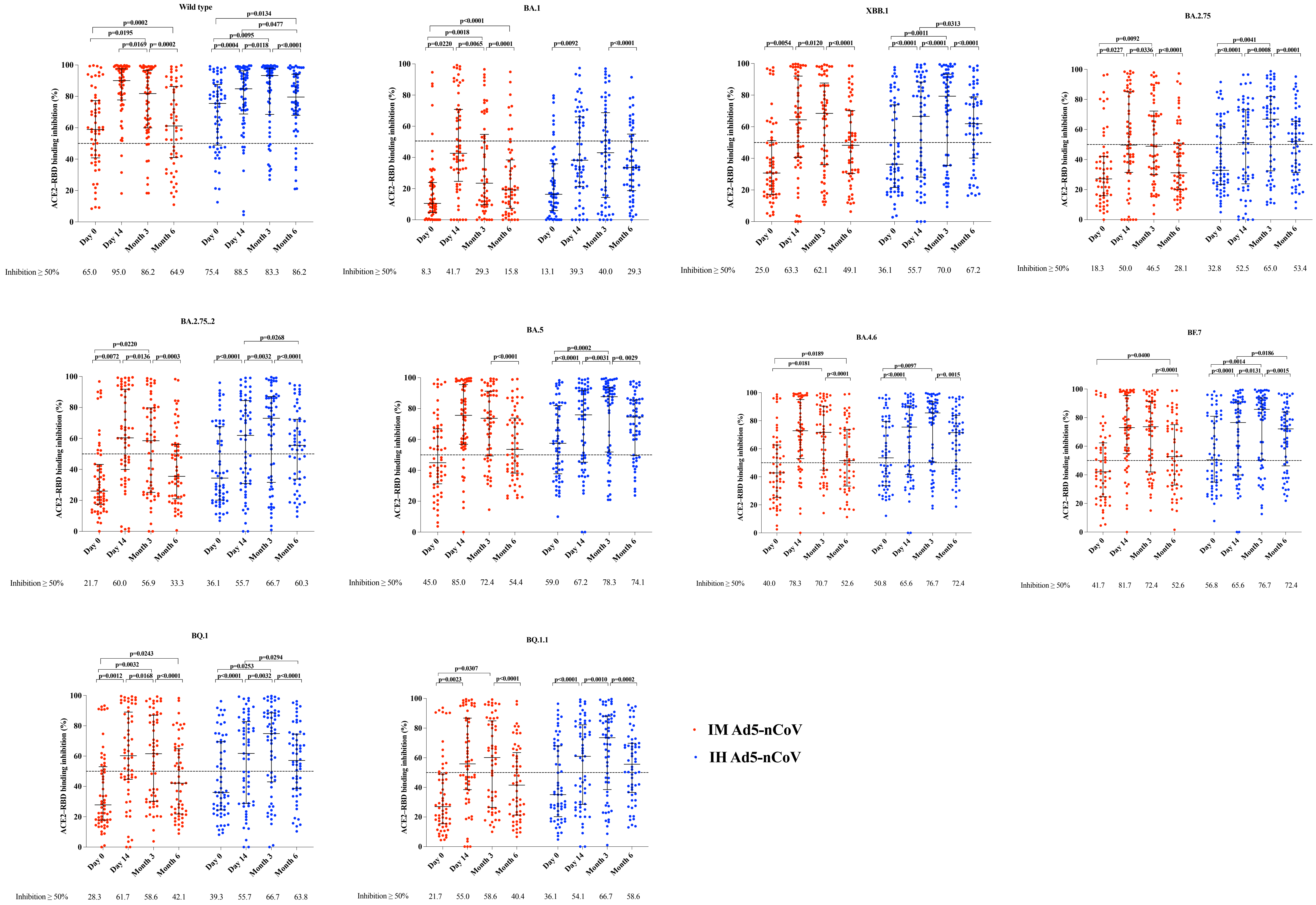

**Figure S4: Specific T-cell responses measured by ELISpot before and after a booster vaccination**

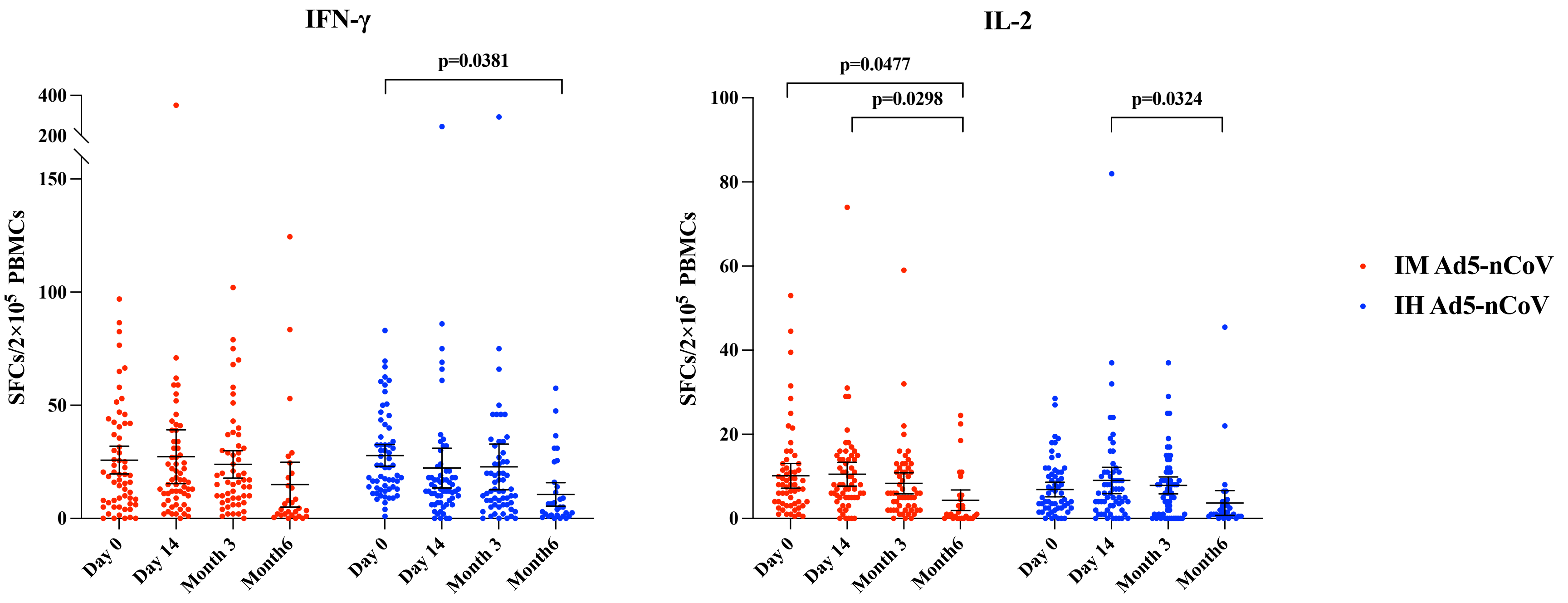

IFN=interferon. IL=interleukin. SFCs=Spot forming cells. PBMCs=peripheral blood mononuclear cells.

**Table S1: Incidence of serious adverse events (SAEs) within 6 months after vaccination classified by system organ class in full analysis population**

| SAEs within 6 months | IM Ad5-nCoV (n=2050) | IH Ad5-nCoV (n=2039) | Control (n=2008) | IM Ad5-nCoV vs. control p value | IM Ad5-nCoV vs. control p value |
|---|---|---|---|---|---|
| **Any Event** | 9 (0.44, 0.23 to 0.83) | 7 (0.34, 0.17 to 0.1) | 6 (0.30, 0.14 to 0.65) | 0.4618 | 0.8025 |
| Renal and urinary disorders | 1 (0.05, 0.01 to 0.28) | 0 | 1 (0.05, 0.01 to 0.28) | > 0.9999 | 0.4962 |
| Endocrine disorders | 1 (0.05, 0.01 to 0.28) | 0 | 1 (0.05, 0.01 to 0.28) | > 0.9999 | 0.4962 |
| Respiratory, thoracic and mediastinal disorders | 3 (0.14, 0.05 to 0.43) | 3 (0.15, 0.05 to 0.43) | 1 (0.05, 0.01 to 0.28) | 0.6249 | 0.6249 |
| Musculoskeletal and connective tissue disorders | 1 (0.05, 0.01 to 0.28) | 0 | 1 (0.05, 0.01 to 0.28) | > 0.9999 | 0.4962 |
| Nervous system disorders | 1 (0.05, 0.01 to 0.28) | 2 (0.10, 0.03 to 0.36) | 1 (0.05, 0.01 to 0.28) | > 0.9999 | > 0.9999 |
| Cardiac disorders | 1 (0.05, 0.01 to 0.28) | 0 | 0 | > 0.9999 | > 0.9999 |
| Gastrointestinal disorders | 1 (0.05, 0.01 to 0.28) | 1 (0.05, 0.01 to 0.28) | 0 | > 0.9999 | > 0.9999 |
| Vascular disorders | 0 | 1 (0.05, 0.01 to 0.28) | 1 (0.05, 0.01 to 0.28) | 0.4948 | > 0.9999 |
| Immune system disorders | 0 | 1 (0.05,0.01 to 0.28) | 0 | > 0.9999 | > 0.9999 |

Data are n (%, 95%CI).

**Table S2: List of serious adverse events (SAEs) within 6 months after vaccination in full analysis population**

| Group | Subject ID | SAE Term | System Organ Class | Preferred Term | Onset date | End date | Duration | Days to onset date from last dose | Grade | Outcome | Causality assessment |
|---|---|---|---|---|---|---|---|---|---|---|---|
| IM Ad5-nCoV | S01-27084 | Renal calculi | Renal and urinary disorders | Nephrolithiasis | 2024-01-29 | 2024-02-01 | 3 | 165 | 3 | Recovered | Unrelated |
| IM Ad5-nCoV | S01-28037 | Malignant thyroid tumor | Endocrine disorders | Thyroid cancer | 2024-01-18 | 2024-01-24 | 6 | 147 | 3 | Relieved | Unrelated |
| IM Ad5-nCoV | S01-50009 | Pulmonary tuberculosis | Respiratory, thoracic and mediastinal disorders | Pulmonary tuberculosis | 2023-11-13 | NA | NA | 150 | 3 | Relieved | Unrelated |
| IM Ad5-nCoV | S01-50022 | Right femoral neck fracture | Musculoskeletal and connective tissue disorders | Femoral neck fracture | 2023-11-05 | 2023-11-18 | 13 | 114 | 3 | Recovered | Unrelated |
| IM Ad5-nCoV | S04-00399 | Pulmonary infection | Respiratory, thoracic and mediastinal disorders | Respiratory tract infection | 2023-12-08 | 2023-12-16 | 8 | 172 | 3 | Recovered | Unrelated |
| IM Ad5-nCoV | S04-00548 | Posterior circulation ischemia | Nervous system disorders | Vertebrobasilar insufficiency | 2023-10-05 | 2023-10-10 | 5 | 107 | 3 | Relieved | Unrelated |
| IM Ad5-nCoV | S04-01531 | Paroxysmal supraventricular tachycardia | Cardiac disorders | Supraventricular tachycardia | 2023-08-13 | 2023-08-23 | 10 | 40 | 3 | Relieved | Unrelated |
| IM Ad5-nCoV | S05-01447 | Community acquired pneumonia | Respiratory, thoracic and mediastinal disorders | Pneumonia | 2023-08-08 | 2023-09-15 | 38 | 26 | 3 | Recovered | Unrelated |
| IM Ad5-nCoV | S05-01996 | Abdominal pain | Gastrointestinal disorders | Abdominal pain | 2023-09-15 | 2023-10-09 | 24 | 57 | 3 | Recovered | Unrelated |
| IH Ad5-nCoV | S04-00162 | Vestibular neuritis | Nervous system disorders | Vestibular neuronitis | 2023-11-17 | 2023-11-24 | 7 | 157 | 3 | Relieved | Unrelated |
| IH Ad5-nCoV | S04-00361 | Pulmonary infection | Respiratory, thoracic and mediastinal disorders | Respiratory tract infection | 2023-06-28 | 2023-07-10 | 12 | 9 | 3 | Recovered | Unrelated |
| IH Ad5-nCoV | S02-01512 | Pulmonary nodule | Respiratory, thoracic and mediastinal disorders | Pulmonary mass | 2023-12-01 | 2023-12-15 | 14 | 119 | 3 | Relieved | Unrelated |
| IH Ad5-nCoV | S03-00164 | Acute allergic reaction | Immune system disorders | Hypersensitivity | 2023-06-02 | 2023-06-03 | 1 | 0 | 3 | Relieved | Related |
| IH Ad5-nCoV | S03-00164 | Hypertension | Vascular disorders | Hypertension | 2023-06-04 | 2023-06-27 | 23 | 2 | 3 | Relieved | Unrelated |
| IH Ad5-nCoV | S03-00164 | Dizziness | Nervous system disorders | Dizziness | 2023-09-30 | 2023-10-06 | 6 | 120 | 3 | Relieved | Unrelated |
| IH Ad5-nCoV | S03-00164 | Hypertension | Vascular disorders | Hypertension | 2023-11-11 | 2023-11-15 | 4 | 162 | 3 | Relieved | Unrelated |
| IH Ad5-nCoV | S04-01212 | Cerebral infarction | Nervous system disorders | Cerebral infarction | 2023-07-10 | 2023-07-18 | 8 | 13 | 3 | Relieved | Unrelated |
| IH Ad5-nCoV | S05-01786 | Bronchitis | Respiratory, thoracic and mediastinal disorders | Bronchitis | 2023-12-20 | 2024-01-18 | 29 | 156 | 3 | Relieved | Unrelated |
| IH Ad5-nCoV | S04-01573 | Gastrointestinal bleeding | Gastrointestinal disorders | Gastrointestinal haemorrhage | 2023-12-19 | 2023-12-21 | 2 | 168 | 3 | Recovered | Unrelated |
| Control | S04-01718 | Type 2 diabetes mellitus with uncontrolled blood sugar | Endocrine disorders | Type 2 diabetes mellitus | 2023-10-22 | 2023-11-03 | 12 | 110 | 3 | Relieved | Unrelated |
| Control | S04-01726 | Cerebral infarction recovery period | Nervous system disorders | Cerebral infarction | 2024-01-02 | 2024-01-29 | 27 | 182 | 3 | Relieved | Unrelated |
| Control | S04-01930 | Lumbar disc herniation | Musculoskeletal and connective tissue disorders | Intervertebral disc protrusion | 2023-12-21 | 2023-12-30 | 9 | 140 | 3 | Relieved | Unrelated |

| | | | | | | | | | | | |
|---|---|---|---|---|---|---|---|---|---|---|---|
| Control | S04-01956 | Chronic kidney disease stage 5 | Renal and urinary disorders | End stage renal disease | 2023-12-12 | 2023-12-22 | 10 | 131 | 3 | Relieved | Unrelated |
| Control | S04-02013 | Coronary artery atherosclerotic heart disease | Vascular disorders | Arteriosclerosis | 2023-11-02 | 2023-11-10 | 8 | 91 | 3 | Relieved | Unrelated |
| Control | S04-02036 | Pulmonary abscess | Respiratory, thoracic and mediastinal disorders | Lung abscess | 2024-01-14 | 2024-03-15 | 61 | 164 | 3 | Recovered | Unrelated |
