## Appendix 2 for "Effectiveness of a booster dose of aerosolized or intramuscular adenovirus type 5 vectored COVID-19 vaccine in adults with hybrid immunity against COVID-19: a multicenter, partially randomized, platform trial in China"

Effectiveness of a COVID-19 booster vaccination in people with hybrid immunity

**Official Title**: Effectiveness of a COVID-19 booster vaccination in Chinese adults aged 18 years or above with hybrid immunity against COVID-19 obtained from both vaccination with COVID-19 vaccines and breakthrough infection of SARS-CoV-2: a multicenter, partially randomized, platform trial

**Sponsor**: Jiangsu Provincial Center for Disease Control and Prevention

Version number: 3.0

Version date: May 17, 2023

| Brief Title | Effectiveness of a COVID-19 booster vaccination in people with hybrid immunity |
| --- | --- |
| Official Title | Effectiveness of a COVID-19 booster vaccination in Chinese adults aged 18 years or above with hybrid immunity against COVID-19 obtained from both vaccination with COVID-19 vaccines and breakthrough infection of SARS-CoV-2: a multicenter, partially randomized, platform trial |
| Version date | May 17, 2023 |
| Version number | 3.0 |

| **Principle Investigator** | Jing-Xin Li | Chief physician | Jiangsu Provincial Center for Disease Control and Prevention |
| --- | --- | --- | --- |
| **Leading Authors** | Feng-Cai Zhu | Chief physician | Jiangsu Provincial Center for Disease Control and Prevention |
|  | Jing-Xin Li | Chief physician | Jiangsu Provincial Center for Disease Control and Prevention |
|  | Zhi-Guo Wang | Chief physician | Jiangsu Provincial Center for Disease Control and Prevention |
|  | Yuan-Bao Liu | Chief physician | Jiangsu Provincial Center for Disease Control and Prevention |
|  | Yang Zhao | Professor | Nanjing Medical University |
|  | Si-Yue Jia | Physician in charge | Jiangsu Provincial Center for Disease Control and Prevention |
|  | Xue-Wen Wang | Senior statistician | Shanghai Canming Medical Technology Co., Ltd |

Copyright 2023 Unauthorized reproduction or use is prohibited

### DOCUMENT HISTORY

| **Version No.** | | **Version Date** | | **Amendment** | |
| --- | --- | --- | --- | --- | --- |
| 1.0 | | March 23 | | N/A | |
| 2.0 | | April 17 | | 1^st^ Amendment | |
| 3.0 | | May 17 | | 2^nd^ Amendment | |
| **Information of the 1^st^ Amendment** | | | | | |
| **Contents in Original Version (1.0)** | | | **Contents in Altered Version (2.0)** | | |
| **Chapter** | **Original Contents** | | **Page/Row** | | **Altered Contents** |
| Chapter 6.1 Target population | The eligible participants aged 18 years and over, including the elderly over 60 years old or those with underlying diseases. Participation is voluntary and an informed consent form is signed. | | **/** | | The eligible participants aged 18 years and over, including the elderly over 60 years old or those with underlying diseases (history of underlying medical conditions diagnosed by a clinician, including hypertension, diabetes, heart disease, etc). Participation is voluntary and an informed consent form is signed. |
| Chapter 6.2 Inclusion and exclusion criteria | **/** | | **/** | | 8. Acute disease or acute onset of chronic disease.  9. Epilepsy and other progressive neurological disorders. |
| **Information of the 2^st^ Amendment** | | | | | |
| **Contents in Original Version (2.0)** | | | **Contents in Altered Version (3.0)** | | |
| **Chapter** | **Original Contents** | | **Page/Row** | | **Altered Contents** |
| Full version | the second COVID-19 vaccine booster | | / | | The first or second COVID-19 vaccine booster |
| Chapter 6.2  Inclusion and exclusion criteria | 3. ≥ 4 months from the last SARS-CoV-2 infection (or never been infected), and 6 months or more from the first booster immunization of the COVID-19 vaccine. | | / | | 3. ≥ 4 months from the last SARS-CoV-2 infection (or never been infected), and 6 months or more from the prime immunization of the COVID-19 vaccine. |
| Investigational vaccine | Vaccine 1：Intramuscular adenovirus type 5 vectored COVID-19 vaccine  Vaccine 2：Aerosolized adenovirus type 5 vectored COVID-19 vaccine  Vaccine 3：DelNS1-2019-nCoV-RBD-OPT1  Vaccine 4：SYS6006 | | / | | Vaccine 1：Intramuscular adenovirus type 5 vectored COVID-19 vaccine  Vaccine 2：Aerosolized adenovirus type 5 vectored COVID-19 vaccine |

**Principal Investigator Statement**

| **Brief Title** | Effectiveness of a COVID-19 booster vaccination in people with hybrid immunity |
| --- | --- |
| **Version number** | 3.0 |
| **Version date** | May 17, 2023 |

I agree with：

- Responsible for correctly guiding the conduct of clinical research in the region.
- Ensure that this trial is conducted in accordance with the trial protocol and clinical research standard operating procedures.
- Ensure that personnel involved in this project fully understand the product information and other research-related responsibilities and obligations specified in this trial protocol.
- Ensure that no changes to the trial protocol are made without review and written approval by the Ethics Committee (IEC), unless necessary to eliminate immediate harm to participants or to comply with the requirements of the registration authority (e.g. project administration) management).
- I am familiar with and will comply with the Good Clinical Practice (GCP), the Guiding Principles for Good Clinical Trial of Vaccines (Trial) and all current regulatory requirements.

**Name of Principal investigator：**Jing-Xin Li

**Signature of principal investigator：**Jing-Xin Li

**Date：**May 17, 2023

**PROTOCOL SUMMARY**

| **Brief Title** | Effectiveness of a COVID-19 booster vaccination in people with hybrid immunity |
| --- | --- |
| **Official Title** | Effectiveness of a COVID-19 booster vaccination in Chinese adults aged 18 years or above with hybrid immunity against COVID-19 obtained from both vaccination with COVID-19 vaccines and breakthrough infection of SARS-CoV-2: a multicenter, partially randomized, platform trial |
| **Objectives** | **Primary objective:**  To evaluate the effectiveness of the COVID-19 booster vaccination in preventing virologically confirmed (positive antigen rapid test or nucleic acid test) COVID-19 disease in people aged 18 years and above with hybrid immunity.  **Secondary objective:**  To evaluate the safety and immunogenicity of the COVID-19 booster immunization in people aged 18 years and above with hybrid immunity. |
| **Target disease** | To prevent COVID-19 caused by SARS-CoV-2 |
| **Target population** | Adults aged 18 years and above, including the elderly aged 60 years and above. |
| **Sample size** | About 6,000 participants. |
| **Rational and background** | On December 13, 2022, the Comprehensive Group of the Joint Prevention and Control Mechanism of the State Council in response to COVID-19 epidemic issued the “Notice on Issuing the Implementation Plan for the Second Booster Immunization of the COVID-19 Vaccine”. The notice clarified: at this stage, on the basis of the first dose of booster immunization, the second dose booster can be carried out among people at high risk of infection, the elderly over 60 years of age, people with serious underlying diseases and low immunity. According to the progress of vaccine development, all vaccines approved for conditional marketing or emergency use can be used for the second dose of booster immunization. Priority should be given to heterologous booster vaccination, or a second dose booster with a vaccine containing the Omicron strain or with good cross-immunity against the Omicron strain. According to worldwide real-world research and clinical trial data, combined with the actual vaccination situation in China, the time interval between the second and first booster dose is more than 6 months. However, there is currently no data on the large-scale protective efficacy of the COVID-19 booster vaccination for people with hybrid immunity. The aim of this study is to conduct a multicenter, partially randomized, platform trial to evaluate the effectiveness of the COVID-19 booster immunization in individuals with hybrid immunity. |
| **Investigational vaccine** | Vaccine 1：Intramuscular adenovirus type 5 vectored COVID-19 vaccine  Vaccine 2：Aerosolized adenovirus type 5 vectored COVID-19 vaccine  Both vaccines are manufactured by CanSino Biologics |
| **Trial design** | This is a multicenter, partially randomized, platform trial aims to evaluate the effectiveness of a booster dose of aerosolized or intramuscular adenovirus type 5 vectored COVID-19 vaccine through a six-month surveillance of COVID-19 in adults aged 18 years and older with hybrid immunity against COVID-19. Eligible participants were individuals aged 18 years and older, including those over 60 years of age and individuals with underlying diseases. Eligibility required an interval ≥ 4 months after previous SARS-CoV-2 infection or confirmation of never having been infected, and ≥ 6 months since the last COVID-19 vaccination. Participants willing to receive a booster dose were randomly assigned to receive one of the boost vaccines, while those opting not to receive a booster were included as a control group. All participants were monitored for symptom-driven COVID-19 for 6 months after the booster dose (in the vaccine groups) or from the time of enrollment (in the control group). **Sample size calculation:** We hypothesized that participants in the vaccinated group receiving a booster shot would exhibit approximately 50% protection compared to those in the control group who did not receive a booster shot. The cumulative incidence rate of COVID-19 endpoint cases in the control group over the 6-month period of was estimated to be around 5%, while it was projected to be about 2.5% in the vaccine group. Sample size calculation was conducted using group-sequential tests for two proportions via PASS software (version 16.0), applying a one-sided α value of 0.05 and aiming for a statistical power of 90%. This analysis indicated that each vaccine group and the control group would require a minimum of 1308 participants. Anticipating a 30% dropout rate, a target enrollment of 2000 participants per group was established. **Table 2 Sample size** group \| **Sample size（case）** \| **immunogenicity subgroup（case）**; Vaccine group 1 \| 2000 \| 60; Vaccine group 2 \| 2000 \| 60; Control group \| 2000 \| /; **total** \| **6000** \| **120** Note: The first 60 participants in each vaccine group will be included in the immunogenicity subgroup. **Study process：** All participants will be monitored for symptom-driven COVID-19 for 6 months after the booster dose (in the vaccine groups) or from the time of enrollment (in the control group). This surveillance will combine active and passive monitoring strategies. We will provide antigen rapid test kits for SARS-CoV-2 infection to both participants in the vaccine groups or control group. Participants will be instructed to conduct a self-test following the product mannul of the antigen rapid test kits in the event that they exhibited any COVID-19 suspected symptoms during the surveillance period. Participants will be instructed to perform an antigen rapid test within 24 hours after the appearance of the suspected first symptom. In addition, the researchers will conduct telephone visits every seven days to proactively remind participants to perform self-tests with the COVID-19 antigen kit after developing suspected symptoms of respiratory infection. For participants in the vaccine groups, the researcher’s telephone interviews will also collect the occurrence of serious adverse events. Among them, participants in the immunogenicity subgroup will be collected blood samples before the booster dose, at 14 days, 3 months and 6 months post-booster dose, with a volume of approximately 10 ml of blood collected each time for isolation of peripheral blood single nucleated cells and serum. In addition, all participants will be collected oral mucosal samples on the day of enrollment. **Effectiveness monitoring and processing:** COVID-19 suspected symptoms include dry throat, sore throat, cough, fever, muscle aches, decreased or loss of smell and taste, nasal congestion, runny nose, diarrhea, conjunctivitis, fatigue, malaise, headache, dyspnea, and nausea. During the observation period of the study, if a participant develops any of the above symptoms, the participant will be required to undergo an antigen test on the 2nd day after the onset of symptoms. If the initial result is negative, they will be required to conduct additional tests at intervals of at least 24 hours until achieving three consecutive negative results. If the initial result was negative, they were required to conduct additional tests at intervals of at least 24 hours until achieving three consecutive negative results. In cases where a positive antigen test result occurred, participants were mandated to promptly inform the investigators. Subsequently, investigators took a throat swab for the positive cases within 48 hours after receiving the positive rapid test report for nucleic acid test. Investigators followed up on each positive case through weekly telephone consultations until the resolution of symptoms or recovery. Each episode of COVID-19 will be classified as mild, moderate, severe, or critical. **The definition of COVID-19 infection history in this study:** Positive nucleic acid or antigen test results; or no pathogenic test results, but acute onset of two or more of the above typical symptoms or signs, and a history of contact with possible or confirmed cases or simultaneous space and time. **Endpoint case:** COVID-19 endpoint cases are defined as participants with COVID-19 confirmed by positive antigen rapid test or nucleic acid test after receiving the booster dose. **Graded COVID-19：** Clinical COVID-19 grading needs to be carried out on the basis of endpoint cases in accordance with the Diagnostic and Treatment Protocol for Pneumonia with SARS-CoV-2 Infection (Trial 10th Edition). （1）Mild The main symptoms are upper respiratory tract infections, such as dry throat, sore throat, cough, fever, etc. （2）Moderate Persistent high fever for > 3 days or/and cough, shortness of breath, etc., but the respiratory rate (RR) is < 30 times/min, and the oxygen saturation when breathing air at rest is > 93%. Imaging studies show characteristic symptoms of pneumonia caused by COVID-19. （3）Severe Adults meet any of the following conditions and cannot be explained by reasons other than COVID-19 infection: 1. Shortness of breath, RR ≥ 30 times/min; 2. In a resting state, oxygen saturation is ≤ 93% when breathing air; 3. Arterial partial pressure of oxygen (PaO2)/oxygen concentration (FiO2) ≤ 300mmHg (1mmHg=0.133kPa). In high-altitude areas (over 1000 meters above sea level), PaO2/FiO2 should be corrected according to the following formula: PaO2/FiO2× [760/atmosphere (mmHg)]; 4. Clinical symptoms progressively worsen, and lung imaging shows that the lesions progress significantly > 50% within 24 to 48 hours. （4）Critical Those who meet one of the following conditions: 1. Respiratory failure occurs and mechanical ventilation is required; 2. Shock occurs; 3. Combined with other organ failure and require ICU monitoring and treatment. *** *It is possible that the participants may be unable to participate in face-to-face interviews due to their own constraints. This study also allows participants to independently visit a nucleic acid testing institution for a PCR test, and submit the test report to the researcher via photography or mail. At the same time, the researcher will contact the participants to ask their specific circumstances.* |
| **Endpoints** | **Primary endpoint**  **Effectiveness：**  The incidence of COVID-19 endpoint cases from 14 days to 6 months after receiving the booster dose.  **Secondary endpoints**  **Effectiveness：**  1. The incidence of COVID-19 endpoint cases from 7 days to 6 months after receiving the booster dose.  2. The incidence of COVID-19 endpoint cases from 28 days to 6 months after receiving the booster dose.  3. The incidence of graded COVID-19 endpoint cases (mild, moderate, severe, critical or death) from 7 days to 6 months, 14 days to 6 months, and 28 days to 6 months after receiving the booster dose.  4. The incidence of hospitalized COVID-19 endpoint cases from 7 days to 6 months, 4 days to 6 months, and 28 days to 6 months after receiving the booster dose.  **Immunogenicity:**  1. GMT, GMI, and positive conversion rate of neutralizing antibodies against the original strain and Omicron variant strain in the serum of participants in the immunogenicity subgroup on 14 days, 3 months, and 6 months after receiving the booster dose.  2. ACE2-RBD binding inhibition rates against the original strain and Omicron variant strain in the serum of participants in the immunogenicity subgroup on 14 days, 3 months, and 6 months after receiving the booster dose.  3. Specific T-cell responses measured by ELISpot assay of participants in the immunogenicity subgroup on 14 days, 3 months, and 6 months after receiving the booster dose.  **Safety:**  The incidence of serious adverse events within 6 months after receiving the booster dose.  **Exploratory Endpoints**  The impact of the human genome on the effect of vaccination. |
| **Scheduled site visits** | All participants in this study need to undergo an interview at V1, among which the immunogenicity subgroup requires a total of 4 interviews at V1~V4. In addition to the above interview content, the vaccine group will undergo 6-month effectiveness monitoring after completing the booster immunization/the control group will receive 6-month effectiveness monitoring after enrollment. Visits will be conducted remotely via phone calls, WeChat messages, text messages, or emails, a total of 24 remote visits (if the face-to-face visit and remote visit overlap, no remote visit is required). Vaccine group V1: Informed consent, collection of demographic information (age, gender, height, weight), medical history inquiry (including history of COVID-19 infection, clinically diagnosed serious chronic diseases such as hypertension, diabetes, cardiovascular disease, and other immune impairments) damaging diseases or impaired immunity caused by long-term medication), inclusion and exclusion criteria screening, allocation of research numbers, blood collection (only applicable to immunogenicity subgroup), vaccination, and observation for 30 minutes, collect oral specimens and distribute COVID-19 antigen rapid test kits. Control group V1: Informed consent, collection of demographic information (age, gender, height, weight), medical history inquiry (including history of COVID-19 infection, clinically diagnosed serious chronic diseases such as hypertension, diabetes, cardiovascular disease, and other conditions that cause immune impairment) damaging diseases or impaired immunity caused by long-term medication), inclusion and exclusion criteria screening, assigning research numbers, collecting oral specimens, and issuing COVID-19 antigen rapid test kits. V2-V4: Immunogenicity blood collection (immunogenicity subgroup); effectiveness monitoring and SAE monitoring. **Table 3 Interview plan and content** **Visit point** \| **Time (window period)** \| **Content**; V1 \| day-1~0 \| Vaccine group：Informed consent, collection of demographic information (age, gender, height, weight), medical history inquiry (including history of COVID-19 infection, clinically diagnosed serious chronic diseases such as hypertension, diabetes, cardiovascular disease, and other immune impairments) damaging diseases or impaired immunity caused by long-term medication), inclusion and exclusion criteria screening, allocation of research numbers, blood collection (only applicable to immunogenicity subgroup), vaccination, and observation for 30 minutes, collect oral specimens and distribute COVID-19 antigen rapid test kits. Control group V1: Informed consent, collection of demographic information (age, gender, height, weight), medical history inquiry (including history of COVID-19 infection, clinically diagnosed serious chronic diseases such as hypertension, diabetes, cardiovascular disease, and other conditions that cause immune impairment) damaging diseases or impaired immunity caused by long-term medication), inclusion and exclusion criteria screening, assigning research numbers, collecting oral specimens, and issuing COVID-19 antigen rapid test kits.; V2 \| 14 days after the booster vaccination（+3 days） \| Immunogenicity blood collection (immunogenicity subgroup); effectiveness monitoring and SAE monitoring.; V3 \| 3 months after the booster vaccination（+15 days） \| Immunogenicity blood collection (immunogenicity subgroup); effectiveness monitoring and SAE monitoring.; V4 \| 6 months after the booster vaccination（+30 days） \| Immunogenicity blood collection (immunogenicity subgroup); effectiveness monitoring and SAE monitoring. **Table 4** **Immunogenicity subgroup blood collection schedule** **Visit point** \| **V1** \| **V2** \| **V3** \| **V4**; Blood sample \| 10ml anticoagulated blood \| 10ml anticoagulated blood \| 10ml anticoagulated blood \| 10ml anticoagulated blood; Total \| 40ml \| \| \| Note: One month is calculated as 28 days. |
| **Statistic analyses** | Final analysis will be conducted after completion of the 6-month follow-up. |
| **Inclusion criteria** | **Inclusion Criteria:**  1. Adults aged 18 years and over, including the elderly over 60 years and those with underlying diseases.  2. Being able and willing to comply with the requirements of the clinical trial protocol and sign the informed consent form.  3. ≥ 4 months from the last SARS-CoV-2 infection (or never been infected), and 6 months or more from the last dose of COVID-19 vaccine. |
| **Exclusion criteria** | **Exclusion Criteria:**  1. Having suspected symptoms of COVID-19 when enrolled, such as dry throat, sore throat, cough, etc.  2. The COVID-19 antigen rapid test is positive when enrolled.  3. Fever, temperature > 37.0°C.  4. Have received a second COVID-19 booster immunization.  5. Have a history of serious adverse reactions related to the vaccine and/or have a history of severe allergic reactions to any component of the investigational vaccine (e.g., systemic allergic reactions) (only applicable to the vaccine groups).  6. Pregnant or lactating women.  7. HIV infection, tuberculosis, low immunity caused by disease or long-term medication.  8. Acute disease or acute onset of chronic disease.  9. Epilepsy and other progressive neurological disorders.  10. Other situations that are not suitable for participating in this research, according to the judgment of the researcher. |

### 1. BACKGROUND

On December 13, 2022, the Comprehensive Group of the Joint Prevention and Control Mechanism of the State Council in response to COVID-19 epidemic issued the “Notice on Issuing the Implementation Plan for the Second Booster Immunization of the COVID-19 Vaccine”. The notice clarified: at this stage, on the basis of the first dose of booster immunization, the second dose booster can be carried out among people at high risk of infection, the elderly over 60 years of age, people with serious underlying diseases and low immunity. According to the progress of vaccine development, all vaccines approved for conditional marketing or emergency use can be used for the second dose of booster immunization. Priority should be given to heterologous booster vaccination, or a second dose booster with a vaccine containing the Omicron strain or with good cross-immunity against the Omicron strain. According to worldwide real-world research and clinical trial data, combined with the actual vaccination situation in China, the time interval between the second and first booster dose is more than 6 months. However, there is currently no data on the large-scale protective efficacy of the COVID-19 booster vaccination for people with hybrid immunity. The aim of this study is to conduct a multicenter, partially randomized, platform trial to evaluate the effectiveness of the COVID-19 booster immunization in individuals with hybrid immunity.

### 2. RELATED PARTIES IN CLINICAL TRIAL

Responsible Party: Jiangsu Provincial Center for Disease Control and Prevention

Locations: Taizhou Gaogang District, Changzhou Wujin District, Lianyungang Ganyu District, Suqian Sucheng District, Wuxi

Data Management and Statistical Party：Shanghai Canming Medical Technology Co., Ltd

Interim Analysis Independent Statistical Party：School of Public Health Nanjing Medical University.

Testing Party：National Institutes for Food and Drug Control

### 3. OBJECTIVES AND EVALUATION INDEX

#### 3.1 Objectives

Primary objective: To evaluate the effectiveness of the COVID-19 vaccine booster vaccination in preventing virologically confirmed (positive antigen rapid test or nucleic acid test) COVID-19 disease in people aged 18 years and above with hybrid immunity.

Secondary objective: To evaluate the safety and immunogenicity of the COVID-19 vaccine booster immunization for people aged 18 years and above with hybrid immunity.

**3.2 Endpoints**

**3.2.1 Primary endpoint**

**Effectiveness：**

The incidence of COVID-19 endpoint cases from 14 days to 6 months after receiving the booster dose.

**3.2.2 Secondary endpoints**

**Effectiveness：**

1. The incidence of COVID-19 endpoint cases from 7 days to 6 months after receiving the booster dose.

2. The incidence of COVID-19 endpoint cases from 28 days to 6 months after receiving the booster dose.

3. The incidence of graded COVID-19 endpoint cases (mild, moderate, severe, critical or death) from 7 days to 6 months, 14 days to 6 months, and 28 days to 6 months after receiving the booster dose.

4. The incidence of hospitalized COVID-19 endpoint cases from 7 days to 6 months, 4 days to 6 months, and 28 days to 6 months after receiving the booster dose.

**Immunogenicity:**

1. GMT, GMI, and positive conversion rate of neutralizing antibodies against the original strain and Omicron variant strain in the serum of participants in the immunogenicity subgroup on 14 days, 3 months, and 6 months after receiving the booster dose.

2. ACE2-RBD binding inhibition rates against the original strain and Omicron variant strain in the serum of participants in the immunogenicity subgroup on 14 days, 3 months, and 6 months after receiving the booster dose.

3. Specific T-cell responses measured by ELISpot assay of participants in the immunogenicity subgroup on 14 days, 3 months, and 6 months after receiving the booster dose.

**Safety:**

The incidence of serious adverse events within 6 months after receiving the booster dose.

**3.2.3 Exploratory endpoints**

The impact of the human genome on the effect of vaccination.

### 4. TRIAL DESIGN

#### 4.1 Overall design

This is a multicenter, partially randomized, platform trial aims to evaluate the effectiveness of a booster dose of aerosolized or intramuscular adenovirus type 5 vectored COVID-19 vaccine through a six-month surveillance of COVID-19 in adults aged 18 years and older with hybrid immunity against COVID-19. Eligible participants were individuals aged 18 years and older, including those over 60 years of age and individuals with underlying diseases. Eligibility required an interval ≥ 4 months after previous SARS-CoV-2 infection or confirmation of never having been infected, and ≥ 6 months since the last COVID-19 vaccination. Participants willing to receive a booster dose were randomly assigned to receive one of the boost vaccines, while those opting not to receive a booster were included as a control group. All participants were monitored for symptom-driven COVID-19 for 6 months after the booster dose (in the vaccine groups) or from the time of enrollment (in the control group).

#### 4.2 Scheduled site visits

All participants in this study need to undergo an interview at V1, among which the immunogenicity subgroup requires a total of 4 interviews at V1~V4. In addition to the above interview content, the vaccine group will undergo 6-month effectiveness monitoring after completing the booster immunization/the control group will receive 6-month effectiveness monitoring after enrollment. Visits will be conducted remotely via phone calls, WeChat messages, text messages, or emails, a total of 24 remote visits (if the face-to-face visit and remote visit overlap, no remote visit is required).

Vaccine group V1: Informed consent, collection of demographic information (age, gender, height, weight), medical history inquiry (including history of COVID-19 infection, clinically diagnosed serious chronic diseases such as hypertension, diabetes, cardiovascular disease, and other immune impairments) damaging diseases or impaired immunity caused by long-term medication), inclusion and exclusion criteria screening, allocation of research numbers, blood collection (only applicable to immunogenicity subgroup), vaccination, and observation for 30 minutes, collect oral specimens and distribute COVID-19 antigen rapid test kits.

Control group V1: Informed consent, collection of demographic information (age, gender, height, weight), medical history inquiry (including history of COVID-19 infection, clinically diagnosed serious chronic diseases such as hypertension, diabetes, cardiovascular disease, and other conditions that cause immune impairment) damaging diseases or impaired immunity caused by long-term medication), inclusion and exclusion criteria screening, assigning research numbers, collecting oral specimens, and issuing COVID-19 antigen rapid test kits.

V2-V4: Immunogenicity blood collection (immunogenicity subgroup); effectiveness monitoring and SAE monitoring.

#### 4.3 Sample size

Sample size calculation was conducted using group-sequential tests for two proportions via PASS software (version 16.0), applying a one-sided α value of 0.05 and aiming for a statistical power of 90%. This analysis indicated that each vaccine group and the control group would require a minimum of 1308 participants. Anticipating a 30% dropout rate, a target enrollment of 2000 participants per group was established.

#### 4.4 Randomization and blinding

The study aims to enroll a total of 6,000 participants who are 18 years old and above. This includes 2,000 individuals in each of the two vaccine groups, as well as a shared control group of 2,000 people. Enrolled population should be randomly vaccinated with different types of vaccines in a ratio of 1:2. According to the block randomization method, an independent randomization statistician generates a participant random table through SAS 9.4 or above version, and imports it into the Interactive Web Response System (IWRS) system, which can only be accessed by authorized personnel. Authorized research center personnel can obtain participant grouping information through the IWRS system and use the experimental vaccines in the corresponding group based on the grouping information.

All participants entering screening will be assigned a screening number. For successfully screened participants, randomization will be performed through the IWRS system and a random number will be obtained (the random number will be used as a unique participant ID number).

This study is open design, and neither the participants nor the researcher will be blinded.

### 5. INVESTIGATIONAL VACCINE

The following two vaccines are proposed to be included in this study for entry into the trial platform.

Biological/Vaccine 1：Intramuscularly administered Ad5-nCoV vaccine

This vaccine is produced by CanSino Biologics Inc.

Biological/Vaccine 2：Aerosolized Ad5-nCoV

This vaccine is produced by CanSino Biologics Inc.

**Inoculation routes, immunization doses and immunization procedures:** The vaccines included in this study are all COVID-19 vaccines that have been conditionally approved by the National Medical Products Administration of China or included in emergency use. The vaccination routes, immunization doses and immunization procedures refer to the instructions for each type of vaccine.

### 6. TARGET POPULATION

#### 6.1 Target population selection

The eligible participants aged 18 years and over, including the elderly over 60 years old or those with underlying diseases (history of underlying medical conditions diagnosed by a clinician, including hypertension, diabetes, heart disease, etc). Participation is voluntary and an informed consent form is signed.

#### 6.2 Inclusion and exclusion criteria

**Inclusion criteria:**

1. Adults aged 18 years and over, including the elderly over 60 years and those with underlying diseases.

2. Being able and willing to comply with the requirements of the clinical trial protocol and sign the informed consent form.

3. ≥ 4 months from the last SARS-CoV-2 infection (or never been infected), and 6 months or more from the last dose of COVID-19 vaccine.

**Exclusion criteria:**

1. Having suspected symptoms of COVID-19 when enrolled, such as dry throat, sore throat, cough, etc.

2. The COVID-19 antigen rapid test is positive when enrolled.

3. Fever, temperature > 37.0°C.

4. Have received a second COVID-19 booster immunization.

5. Have a history of serious adverse reactions related to the vaccine and/or have a history of severe allergic reactions to any component of the investigational vaccine (e.g., systemic allergic reactions) (only applicable to the vaccine groups).

6. Pregnant or lactating women.

7. HIV infection, tuberculosis, low immunity caused by disease or long-term medication.

8. Acute disease or acute onset of chronic disease.

9. Epilepsy and other progressive neurological disorders.

10. Other situations that are not suitable for participating in this research, according to the judgment of the researcher.

#### 6.3 Early withdrawal criteria

Early withdrawal means that the participant fails to complete vaccination, blood collection or follow-up in accordance with the clinical trial protocol, and the researcher decides whether to continue subsequent related studies based on the situation.

Participants can withdraw from the trial at any time according to their own wishes;

Those who leave the observation area and are unable to complete then study process will be deemed to have voluntarily withdrawn from the test.

Intolerable adverse events/SAEs, whether or not related to the investigational vaccine;

The participant’s health condition does not allow him or her to continue to participate in this clinical trial;

Participant withdraws informed consent (data before withdrawing informed consent can still be used);

Other reasons the researchers believe.

### 7. METHODS AND PROCEDURES

#### 7.1 Participant screening

The investigator will conduct a general examination of the volunteers, such as height and weight. The researcher will ask the volunteers according to the inclusion and exclusion criteria of the protocol, and based on the results of the inquiry, decide whether the volunteers will be enrolled in this clinical trial. The physical examination and inclusion and exclusion information of the volunteers will be entered into the vaccination and visit records.

#### 7.2 Enrollment

Screened participants will be given a booster immunization.

#### 7.3 Pre-vaccination sample collection

The participants in the immunogenicity subgroup were also required to undergo blood collection before booster immunization, with a volume of approximately 10 ml of blood collected each time for isolation of peripheral blood single nucleated cells and serum. All participants had oral specimens collected once on the day of enrollment.

#### 7.4 Vaccination

The vaccination routes, immunization doses and immunization procedures refer to the instructions for each type of vaccine.

#### 7.5 On-Site medical observation

Participants need to be under on-site medical observation for 30 minutes after vaccination.

#### 7.6 Post-vaccination sample collection

The participants in the immunogenicity subgroup were also required to undergo additional blood collection on day 0, day 14, 3 months, and 6 months of booster immunization, with a volume of approximately 10 ml of blood collected each time for isolation of peripheral blood single nucleated cells and serum.

#### 7.7 Reporting of serious adverse events

Serious adverse event (SAE), an adverse medical event such as death, life-threatening, permanent or severe disability or loss of function, participant requiring hospitalization or prolonged hospitalization, and congenital anomalies or birth defects that occur after a participant receives an investigational drug.

Upon being informed of the SAE, the investigator should promptly (within 24 hours) report it to the investigator.

Upon receipt of information related to the safety of the vaccine from any source, the investigator should analyze and evaluate the information, including the severity, relevance to the study, and whether it is an anticipated adverse event.

For suspected and unanticipated serious adverse reactions that are fatal or life-threatening, the investigator shall report to the State Drug Administration as soon as possible after the first notification, but not more than 7 natural days, and report the relevant follow-up information within the following 8 days; for suspected and unanticipated serious adverse reactions that are not fatal or life-threatening, or for information on other potential serious safety risks, the investigator shall report to the State Drug Administration as soon as possible after the first notification, but not more than 15 natural days.

#### 7.8 Effectiveness evaluation

All participants will be monitored for symptom-driven COVID-19 for 6 months after the booster dose (in the vaccine groups) or from the time of enrollment (in the control group). This surveillance will combine active and passive monitoring strategies. We will provide antigen rapid test kits for SARS-CoV-2 infection to both participants in the vaccine groups or control group. Participants will be instructed to conduct a self-test following the product mannul of the antigen rapid test kits in the event that they exhibited any COVID-19 suspected symptoms during the surveillance period. Participants will be instructed to perform an antigen rapid test within 24 hours after the appearance of the suspected first symptom. In addition, the researchers will conduct telephone visits every seven days to proactively remind participants to perform self-tests with the COVID-19 antigen kit after developing suspected symptoms of respiratory infection. For participants in the vaccine groups, the researcher’s telephone interviews will also collect the occurrence of serious adverse events. Among them, participants in the immunogenicity subgroup will be collected blood samples before the booster dose, at 14 days, 3 months and 6 months post-booster dose, with a volume of approximately 10 ml of blood collected each time for isolation of peripheral blood single nucleated cells and serum. In addition, all participants will be collected oral mucosal samples on the day of enrollment.

**Effectiveness monitoring and processing:**

COVID-19 suspected symptoms include dry throat, sore throat, cough, fever, muscle aches, decreased or loss of smell and taste, nasal congestion, runny nose, diarrhea, conjunctivitis, fatigue, malaise, headache, dyspnea, and nausea. During the observation period of the study, if a participant develops any of the above symptoms, the participant will be required to undergo an antigen test on the 2nd day after the onset of symptoms. If the initial result is negative, they will be required to conduct additional tests at intervals of at least 24 hours until achieving three consecutive negative results. If the initial result was negative, they were required to conduct additional tests at intervals of at least 24 hours until achieving three consecutive negative results. In cases where a positive antigen test result occurred, participants were mandated to promptly inform the investigators. Subsequently, investigators took a throat swab for the positive cases within 48 hours after receiving the positive rapid test report for nucleic acid test. Investigators followed up on each positive case through weekly telephone consultations until the resolution of symptoms or recovery. Each episode of COVID-19 will be classified as mild, moderate, severe, or critical.

**The definition of COVID-19 infection history in this study:**

Positive nucleic acid or antigen test results; or no pathogenic test results, but acute onset of two or more of the above typical symptoms or signs, and a history of contact with possible or confirmed cases or simultaneous space and time.

**Endpoint case:**

COVID-19 endpoint cases are defined as participants with COVID-19 confirmed by positive antigen rapid test or nucleic acid test after receiving the booster dose.

**Graded COVID-19：**

Clinical COVID-19 grading needs to be carried out on the basis of endpoint cases in accordance with the Diagnostic and Treatment Protocol for Pneumonia with SARS-CoV-2 Infection (Trial 10th Edition).

（1）Mild

The main symptoms are upper respiratory tract infections, such as dry throat, sore throat, cough, fever, etc.

（2）Moderate

Persistent high fever for > 3 days or/and cough, shortness of breath, etc., but the respiratory rate (RR) is < 30 times/min, and the oxygen saturation when breathing air at rest is > 93%. Imaging studies show characteristic symptoms of pneumonia caused by COVID-19.

（3）Severe

Adults meet any of the following conditions and cannot be explained by reasons other than COVID-19 infection:

1. Shortness of breath, RR ≥ 30 times/min;

2. In a resting state, oxygen saturation is ≤ 93% when breathing air;

3. Arterial partial pressure of oxygen (PaO2)/oxygen concentration (FiO2) ≤ 300mmHg (1mmHg=0.133kPa). In high-altitude areas (over 1000 meters above sea level), PaO2/FiO2 should be corrected according to the following formula: PaO2/FiO2× [760/atmosphere (mmHg)];

4. Clinical symptoms progressively worsen, and lung imaging shows that the lesions progress significantly > 50% within 24 to 48 hours.

（4）Critical

Those who meet one of the following conditions:

1. Respiratory failure occurs and mechanical ventilation is required;

2. Shock occurs;

3. Combined with other organ failure and require ICU monitoring and treatment.

*** *It is possible that the participants may be unable to participate in face-to-face interviews due to their own constraints. This study also allows participants to independently visit a nucleic acid testing institution for a PCR test, and submit the test report to the researcher via photography or mail. At the same time, the researcher will contact the participants to ask their specific circumstances.*

### 8. DATA MANAGEMENT

#### 8.1 eCRF design

The eCRF is designed according to the investigational steps specified in the plan. After the first draft is formed, it needs to be reviewed by the project manager, medical, data and statisticians and other project team members to ensure that it conforms to the plan and complies with relevant laws and regulations, and the version control process needs to be fully recorded.

#### 8.2 eCRF filling guide

The eCRF filling guide is the detailed instructions for filling out each page of the eCRF form and each data point. Ensure that the clinical trial center obtains the eCRF and its filling guide before enrolling participants, and trains relevant staff of the clinical trial center on the eCRF filling and data submission process. This process needs to be archived and recorded.

#### 8.3 eCRF annotation

Annotation eCRF is an annotation for a blank eCRF, recording the location of each data item in the eCRF and its variable name and encoding in the database. All data items in the eCRF requires annotation.

#### 8.4 Database design

The database should be established according to the data set name, variable name, variable type and variable length in the annotated eCRF, and try to follow the structure and settings of the standard database. After the database is established, the database should be tested and can be used only after passing the test.

#### 8.5 Permission assignment

System administrators create accounts based on different roles and grant different permissions.

#### 8.6 EDC Fill in

The researcher should follow the requirements of the study protocol to collect the data from the participants and fill in the EDC accurately, timely, complete and standardized according to the original data with reference to the guidelines for filling in the EDC. Modification of the EDC data must follow the standard operation procedures and keep the traces of modification.

#### 8.7 Dispatch and resolution of challenges

The detailed data verification plan provided by the DM of the data management department will undergo review by medical personnel, statisticians, project managers, etc. If there are no objections, the plan will be signed and confirmed by the data administrator and data manager. Once the data is entered into the EDC system, it will undergo verification through the logic checks (Edit Checks) outlined in the plan. Any questionable data will generate system questions automatically, while data that cannot be flagged by the system will be manually questioned through EDC. Researchers will address and respond to both manual and system questions, correcting any incorrect data if necessary until the queries are resolved. In cases where the answers fail to resolve the queries, the data administrator can raise further questions regarding the specific data point, with all interactions being recorded and saved in the EDC database.

#### 8.8 Data modification and review

The researcher can modify the data after verifying the data, and the modified data should follow the system prompts and fill in the reason for modification in the system. The researcher has the authority to review all final data.

#### 8.9 Medical code

Participant medical history and SAEs in clinical trials should be coded using a standardized dictionary. The standard dictionary generally used is the Medical Dictionary for Regulatory Activities (MedDRA). The coded dataset should clearly document the dictionary and version used for coding.

#### 8.10 Data review session

Before locking the database, prepare the first draft of the data review report and all data lists. The database is finalized by the investigator, data manager, and statistical analyst, and the statistical analysis of the population delineation, verification of the records of serious adverse event reports and treatments, etc., in accordance with the clinical trial protocols. The data review meeting is followed by the finalization of the data management report and population delineation plan, etc.

#### 8.11 Database locking and unlocking

Database locking is an important milestone in the clinical research process. The locking process and time should be clearly documented. Locking revokes the permission to edit the database. Any unauthorized account is prohibited from operating the database.

Try to avoid unlocking the database after it is locked. If modification is really necessary, an application must be submitted, which can only be implemented after discussion and signature confirmation by the researcher and data management personnel, and the reasons for unlocking must be recorded in detail.

### 9. STATISTICAL CONSIDERATIONS

In addition to the investigational plan, a separate Statistical Analysis Plan (SAP) will be written to further describe the details of the statistical analysis content and methods. SAP will be finalized before the database is locked.

#### 9.1 Analysis set

**Full Analysis Set (FAS)**：FAS is based on ITT (intention to treat analysis) principle to determine the participants. All of the participants that receiving 1 dose vaccine, and complete pre-immunization blood collection (for the immunogenicity subgroup only), will be included in the FAS set. Among them, participants who were vaccinated incorrectly were randomly divided into groups for immunogenicity evaluation according to the ITT principle.

**Per Protocol Set (PPS)：**Includes all participants who have not violated the inclusion/exclusion criteria, completed immunization, completed pre-immunization blood collection, and have pre-immunization and post-immunization antibody test results (only applicable to the immunogenicity subgroup).

**Safety Set (SS)：**All vaccinated participants will be included. Among them, if the vaccine number is wrong, according to the ASaT (All Participants as Treated) principle, the safety evaluation will be carried out according to the vaccine group actually received by the participant.

The above set of analyses will be discussed and decided by the principal investigator, statistician and data manager in a data blinded review session before the database is locked.

#### 9.2 Statistical analysis methods

Effectiveness analyses will be done in full analysis population, which include all participants who underwent randomization and either received one dose of the vaccine or were enrolled as part of the control group). The incidence of COVID-19 in the control group was calculated based on all endpoint cases identified from the day following enrollment. Effectiveness estimates were derived using the Cox proportional hazards regression model. Moreover, effectiveness adjusted by age, sex, body mass index (BMI) and SARS-CoV-2 infection history was also calculated. Cumulative incidence data were presented using the Kaplan-Meier method. Immunogenicity analyses were restricted to the immunogenicity subgroup, comprising all participants who received vaccinations and provided blood or nasal mucosa samples after vaccination. Safety analyses were performed on the full analysis population. The χ² test or Fisher’s exact test was used for categorical data. Student’s t test was used for log-transformed antibody titers, and the Wilcoxon rank-sum test for data that were not normally distributed. The antibodies against SARS-CoV-2 were reported as geometric mean titers (GMT) with 95% CIs and the cellular responses were shown as the proportion of positive responders.

#### 9.3 Final analysis

Final analysis will be conducted after completion of the 6-month follow-up.

#### 10.1 Ethical review and approval

The principal investigator should submit the clinical trial protocol and all necessary appendix documents to The Ethics Committee for the initial review as required

-	Clinical trial Protocol (indicate the version number/date)

-	Informed consent (indicate the version number/date)

-	Participant recruitment materials (indicate the version number/date)

-	eCRF example draft (indicate the version number/date)

- Vaccination visit records (indicate the version number/date)

- Principal investigator’s CV

The certificate of approval should be issued to the investigator after getting the approval of the ethics committee.

#### 10.2 Follow-up Auditing

To audit the method of participant recruitment, if the information offered to the Participants or impartial witness was completed, understandable; if the informed consent was offered appropriately, if the SAE was reported in time. If there was SAE occurred on the Participants, they could get immediate medical treatment.

During the research period, the Ethics Committee should monitor that if the ratio of risk and benefit increased and if the participants’ rights and interests are effectively protected.

#### 10.3 Potential danger and danger minimization

**10.3.1** **Benefit and Risk**

The participants in this study will not pay for the investigational vaccines. Participants in this clinical trial will receive a second booster immunization of a COVID-19 vaccine that has been conditionally approved or included for emergency use. The participants might be protected against COVID-19 caused by SARS-CoV-2 infection in a period of time after vaccination. At the same time, there may be some adverse reactions following injection. Common vaccination adverse reactions include: fever, tenderness and swelling on the injection site, redness. The adverse reactions are usually relieved in the 3-5 days after they occur.

**10.3.2 Blood Sample collection**

Venous blood samples should be collected by experienced nurses who have gotten trained in accordance with the procedures after the qualification audit of the primary investigator to minimize the pain or danger of participants (including pain and venous puncture site infection which is not common).
